# Cigarette Smoking-Associated Isoform Switching and 3’ UTR Lengthening Via Alternative Polyadenylation

**DOI:** 10.1101/2021.06.09.21258495

**Authors:** Zhonghui Xu, John Platig, Sool Lee, Adel Boueiz, Rob Chase, Dhawal Jain, Andrew Gregory, Rahul Suryadevara, Seth Berman, Russell Bowler, Craig P. Hersh, Alain Laederach, Peter J. Castaldi, for the COPDGene Investigators

## Abstract

**Background:** Cigarette smoking accounts for approximately one in five deaths in the United States. Previous genomic studies have primarily focused on gene level differential expression to identify related molecular signatures and pathways, but the genome-wide effects of smoking on alternative isoform regulation and posttranscriptional modulation have not yet been described.

**Results:** We conducted RNA sequencing (RNA-seq) in whole-blood samples of 454 current and 767 former smokers in COPDGene Study. We assessed the association of current smoking with differential expression of genes and isoforms and differential usage of isoforms and exons. At 10% FDR, we detected 3,167 differentially expressed genes, 2,014 differentially expressed isoforms, 945 differentially used isoforms and 160 differentially used exons. Genes containing differentially used isoforms were enriched in biological pathways involving GTPase activity and innate immunity. The majority of these genes were not differentially expressed, thus not identifiable from conventional differential gene expression analysis. Isoform switch analysis revealed for the first time widespread 3′ UTR lengthening associated with cigarette smoking, where current smokers were found to have higher expression and usage of isoforms with markedly longer 3′ UTRs. The lengthening of 3′ UTRs appears to be mediated through alternative usage of distal polyadenylation sites, and these extended 3′ UTR regions are significantly enriched with functional sequence elements including adenylate-uridylate (AU)-rich elements, microRNA and RNA-protein binding sites. Expression quantitative trait locus analyses on differentially used 3′ UTRs identified 79 known GWAS variants associated with multiple smoking-related human diseases and traits.

**Conclusions:** Smoking elicits widespread transcriptional and posttranscriptional alterations with disease implications. It induces alternative polyadenylation (APA) events resulting in a switch towards the usage of isoforms with strikingly longer 3′ UTRs in genes related to multiple biological pathways including GTPase activity and innate immunity. The extended 3′ UTR regions are enriched with functional sequence elements facilitating post-transcriptional regulation of protein expression and mRNA stability. These findings warrant further studies on APA events as potential biomarkers and novel therapeutic targets for smoking-related diseases.

## Introduction

Cigarette smoking is a major risk factor for a wide range of diseases including cancers, cardiovascular and respiratory diseases. Approximately one in five deaths in the United States is attributable to smoking^1–5^. Globally, smoking-related annual mortality is projected to rise from 3 million in 1995 to 10 million by 2030, with 70% of these deaths occurring in developing countries^2^. The associated socioeconomic burden is enormous, with the proportion of health care expenditure in the US attributable to smoking estimated to range between 6% and 18% across different states^6^.

Smoking cessation has been shown to reverse many smoking-related adverse health effects and substantially reduce mortality^2,7^. At the molecular level, the majority of smoking-deregulated genes revert to normal expression levels following smoking cessation, while a smaller subset of genes remain persistently altered in former smokers^8,9^. While these genomic studies shed light on smoking-related transcriptional modulations at the gene level, few studies have investigated the effect of smoking on alternative isoform regulation. Most multi-exon human genes are expressed in multiple transcript isoforms, and alternative expression of these isoforms are modulated through multiple mechanisms including alternative splicing, alternative promoter usage and alternative polyadenylation. With regulatory impacts on mRNA and protein localization, stability and functional interactions, alternative isoform regulation plays an important role in tissue and cell type specificity and disease susceptibility^10–13^.

In a previous RNA-seq analysis of 515 current and former smokers, we identified instances of differential exon usage predominantly localized to the first or last exons of the involved transcripts, indicating smoking-related alterations in transcription initiation or termination^14^. In the current study, we characterized alternative isoform regulation and associated biological pathways in response to cigarette smoking in a larger RNA-seq sample of 1,221 current and former smokers in the COPDGene Study. We quantified transcriptomic alterations at the gene, isoform and exon level, and analyzed the consequences of alternative isoform usage (i.e. isoform switching^15^). We discovered a widespread switch in current smokers toward increased usage of isoforms with markedly longer 3′ UTRs. This was mediated through alternative usage of distal polyadenylation sites and resulted in the acquisition of additional binding sites for microRNAs (miRNAs) and other functional elements.

## Methods

### Study subjects

This study includes 454 current smokers and 767 former smokers from COPDGene Study^16^. Self-identified non-Hispanic whites and African Americans between the ages of 45 and 80 years with a minimum of 10 pack-years lifetime smoking history were enrolled at 21 centers across the United States. COPDGene conducted two study visits approximately five years apart, and additional longitudinal follow-up of this cohort is ongoing. At the second study visit, complete blood count (CBC) data and PaxGene RNA tubes were collected. Smoking history was ascertained by self-report. Participants defined as current smokers answered yes to the question “Do you smoke cigarettes now (as of one month ago?)”, and for a subset of subjects smoking status was confirmed by serum cotinine measurement. Institutional review board approval and written informed consent was obtained for all subjects.

### Cotinine measurement

Cotinine measurements were obtained from plasma samples of subjects in two COPDGene clinical centers (National Jewish Health and University of Iowa). Plasma was collected using an 8.5 mL p100 tube (Becton Dickinson), and global metabolite data was generated using the Metabolon Global Metabolomics Platform (Durham, NC, USA). The data were normalized to remove batch effects^17^.

### RNA extraction, sequencing and expression quantification

Total RNA was extracted from peripheral blood samples, and paired end reads were generated from Illumina sequencers and aligned to the GRCh38 genome (Supplementary Text 1). GTF annotation was downloaded from Biomart Ensembl database (Ensembl Genes release 94, GRCh38.p12 assembly) on October 21, 2018. Exons from the GTF were broken into disjoint parts (exonic parts) sharing a common set of transcripts^18^. Sequencing read counts on genes and exonic parts were generated from featureCounts in Rsubread^19^ (v1.32.2). Isoform expression estimates were obtained using Salmon^20^ (v0.12.0) and tximport^21^ (v1.10.0).

### Filtering, normalization, differential expression and usage analysis

Low expressed genomic features were filtered before applying TMM^22^ normalization from edgeR^23^ (v3.24.3) (Supplementary Text 1). To test for differential expression of genomic features between current and former smokers, we employed the linear modeling approach implemented in limma^24,25^ (v3.38.3), where the mean-variance relationship is accounted for by applying observation-specific weights estimated from voom^26^. We adjusted for covariates including age, race, gender, total pack-years of exposure, forced expiratory volume in one second (FEV1), complete blood cell count proportions and library prep batch. To test differential usage of isoforms and exonic parts, we used diffSplice from limma. False discovery rate (FDR) was controled with Benjamini-Hochberg procedure^27^. A significance cutoff of 10% FDR was used. To better visualize the differential usage result, we developed a procedure to derive a variance stabilizing transformation (VST) on counts based on the mean-variance relationship in voom (Supplementary Text 2).

### Gene set enrichment analysis

Gene ontology^28,29^ (GO) biological function enrichment of gene sets derived from differential expression and usage analysis were assessed via Fisher exact test statistic with weight01 algorithm available in topGO (v2.33.1) that accounts for dependency in GO topology^30^. P-value < 0.05 was considered significant.

### Isoform switch analysis

Isoforms identified from differential usage analysis were further examined for their splicing patterns relative to a synthetic pre-RNA in their parent genes using IsoformSwitchAnalyzeR (v1.12.0)^31^. Eight categories of splicing events were characterized, including exon skipping (ES), multiple exon skipping (MES), mutually exclusive exons (MEE), intron retention (IR), alternative 5′ splice site (A5), alternative 3′ splice site (A3), alternative transcription start site (ATSS) and alternative transcription termination site (ATTS). By pairwise comparison between down-used and up-used isoforms, we examined eight aspects of isoform switch consequences – namely, changes in overall isoform length, 3′ UTR length, 5′ UTR length, number of exons, intron retention, sensitivity to nonsense-mediated mRNA decay (NMD), location of transcription start site (Tss) and transcription termination site (Tts). The net effects of these splicing events and switch consequences were aggregated at the gene level and tested for statistical significance using a binomial test.

### Sequence and motif analysis

Genomic annotations of polyadenylation cleavage sites (PASs), AU-rich elements (AREs), miRNAs and RNA-binding proteins (RBPs) binding sites were collected from multiple sources (Supplementary Text 1). Flanking sequences of PASs were searched for polyadenylation [poly(A)] signal motifs of AATAAA and TTTTTTTTT. Frequencies of these annotated sequence elements and identified poly(A) signal motifs were computed, smoothed and visualized at each position of a given set of equal-length sequences extracted based on some criterion (e.g. sequences up to 60 nucleotides [nts] upstream of PASs in 3′ UTR exonic parts that were up-used in smokers).

### Statistical, network and eQTL analysis

Demographic differences between current and former smokers were assessed via Student’s t-test and Pearson’s Chi-squared test for continuous and categorical variables, respectively. Isoform and exonic part length comparisons were performed using the Wilcoxon signed rank test. Enrichment tests of sequence elements in 3′ UTRs were performed using Fisher’s exact test. To account for difference in 3′ UTR lengths, we repeated the enrichment analysis limiting to the last 100 nts at 3′ end of the UTRs. To identify individual miRNA and RBPs whose binding sites were enriched at a higher density in 3′ UTRs, we conducted a binomial test with the hypothesized probability of success equal to the ratio of the sum of the lengths of the 3′ UTRs of interest over the total length of 3′ UTRs. The identified individual miRNAs, RBPs and their target genes were visualized as a directed regulatory network using the Fruchterman-Reingold layout^32^, and network communities were detected using a multi-level modularity optimization algorithm implemented in igraph R package (v1.2.5).

Expression quantitative trait locus (eQTL) analyses were performed to test for association between single nucleotide polymorphisms (SNPs) within 1MB *cis* window and the expression values of genes and exonic parts in 796 NHW subjects in COPDGene. SNPs with minor allele frequency > 5% were tested. Expression values were regressed on additively coded SNP genotypes using linear regression implemented in MatrixQTL^33^. The models were also adjusted for age, gender, principal components of genetic ancestry, and 35 PEER factors obtained from the expression data^34^. The identified QTLs at 5% FDR cutoff were cross-referenced against the NHGRI-EBI GWAS catalog accessed on May 07, 2021 using makeCurrentGwascat from gwascat (v2.13.5), and visualized with LocusZoom^35^.

## Data availability

The gene, isoform and exon count data used for this analysis are available in GEO^36,37^ (accession number GSE171730). A Shiny app to explore and visualize the data and result is available at http://cdnm-castaldi.org/smoking_deu_2021/.

## Results

### Differential gene expression

The demographics and clinical characteristics of the study subjects (454 current smokers and 767 former smokers) are summarized in Supplementary Table ST1. In a subset of subjects, serum cotinine levels confirmed the general accuracy of subjects’ self-reported smoking behavior in the COPDGene Study (Supplementary Fig. SF1). To evaluate gene expression changes in peripheral blood in response to active cigarette smoking, we obtained gene level RNA-seq counts, and performed differential gene expression (DGE) analysis comparing current versus former smokers while adjusting for other demographic and clinical covariates. Out of 22,020 genes evaluated, we identified 1,542 up-regulated and 1,625 down-regulated genes at 10% FDR (Supplementary Fig. SF2, Supplementary Table ST2). The top ten DGE genes are listed in Table 1. We then performed GO enrichment analyses on DGE genes and found 335 over-represented biological processes with various aspects of inflammation and platelet activation topping the list (Supplementary Table ST3).

**Table 1.** Top 10 differentially expressed genes in current smokers versus former smokers.

| Ensembl gene ID | HUGO gene name | Chromosome | Strand | Log fold change | Average expression | Adjusted P-value |
| --- | --- | --- | --- | --- | --- | --- |
| ENSG00000154165 | *GPR15* | 3 | + | 1.36 | 4.43 | 3.61E-97 |
| ENSG00000253230 | *LINC00599* | 8 | - | 1.65 | -2.67 | 1.38E-68 |
| ENSG00000173114 | *LRRN3* | 7 | + | 1.17 | 4.19 | 3.36E-55 |
| ENSG00000167680 | *SEMA6B* | 19 | - | 1.35 | -1.16 | 1.01E-47 |
| ENSG00000111961 | *SASH1* | 6 | + | 0.73 | 3.03 | 6.66E-45 |
| ENSG00000063438 | *AHRR* | 5 | + | 1.20 | -0.26 | 3.54E-42 |
| ENSG00000077063 | *CTTNBP2* | 7 | - | 1.30 | -0.70 | 1.59E-41 |
| ENSG00000153823 | *PID1* | 2 | - | 0.51 | 4.03 | 1.10E-39 |
| ENSG00000124334 | *IL9R* | X | + | 0.90 | -0.40 | 1.48E-32 |
| ENSG00000080822 | *CLDND1* | 3 | - | 0.24 | 6.41 | 2.84E-26 |

### Differential expression and usage of isoforms

We next generated Salmon estimates of isoform expression and assessed differential isoform expression (DIE) between current and former smokers. Out of 85,437 isoforms tested, 1,026 up-regulated and 988 down-regulated isoforms were identified at 10% FDR (Supplementary Table ST4, Supplementary Fig. SF3). These isoforms map to 1,547 genes, 77% (1190/1547) of which were also differentially expressed in DGE analysis. The vast majority (1347/1547 = 87%) of these genes had multiple expressed isoforms, and for 64% (860/1347) of these genes the dominant isoform (i.e. most highly expressed isoform) was differentially expressed. GO enrichment analysis identified 290 over-represented biological processes (Supplementary Table ST5), 37% of which were also identified in the DGE enrichment analysis.

Unlike DIE, differential isoform usage (DIU) analysis detects changes in the fractional composition of isoforms originating from the same parent gene (i.e. isoform switch^15^). We identified 389 up-used and 556 down-used isoforms (Supplementary Table ST6), corresponding to 804 genes of which 31% (250/804) were also differentially expressed in the DGE analysis (Supplementary Fig. SF4). Interestingly, DIU occurred largely in non-dominant isoforms (646/804=80%). GO enrichment analysis of genes containing DIU isoforms identified 100 over-represented biological processes (Supplementary Table ST7), 12% of which overlapped with the DGE enrichment results. The most enriched biological processes include GTPase activity, Wnt-signaling, and regulation of innate immunity. The top ten DIU isoforms and enriched GO terms are shown in Tables 2 and 3, respectively.

**Table 2.** Top 10 differentially used Isoforms in current smokers versus former smokers.

| Ensembl transcript ID | HUGO gene name | Log fold change | Average log expression | Adjusted P-value |
| --- | --- | --- | --- | --- |
| ENST00000586582 | *SEMA6B* | 1.54 | -1.41 | 3.04E-21 |
| ENST00000589889 | *SEMA6B* | -1.54 | 0.03 | 3.04E-21 |
| ENST00000233156 | *TFPI* | -0.87 | 0.76 | 3.55E-14 |
| ENST00000244174 | *IL9R* | 0.97 | -1.81 | 1.78E-10 |
| ENST00000540368 | *ATP6V0A2* | -0.77 | -0.19 | 2.66E-10 |
| ENST00000517625 | *SKP1* | -0.43 | 4.11 | 5.83E-10 |
| ENST00000278499 | *SESN3* | -0.61 | 4.19 | 9.34E-10 |
| ENST00000477931 | *GNAS* | -0.56 | 4.13 | 3.76E-09 |
| ENST00000362091 | *FBH1* | -0.44 | 3.94 | 3.76E-09 |
| ENST00000333007 | *TNFAIP2* | -0.87 | 0.48 | 3.80E-09 |

**Table 3.** Top 10 gene ontology biological processes enriched in genes with differentially used isoforms in current smokers versus former smokers.

| GO ID | GO term | Total number of genes in category | Number of smoking-associated genes in category | P-value |
| --- | --- | --- | --- | --- |
| GO:0043547 | positive regulation of GTPase activity | 308 | 45 | 1.00E-05 |
| GO:0046822 | regulation of nucleocytoplasmic transport | 97 | 12 | 3.10E-04 |
| GO:0006607 | NLS-bearing protein import into nucleus | 24 | 7 | 9.50E-04 |
| GO:0010172 | embryonic body morphogenesis | 9 | 5 | 1.27E-03 |
| GO:0008053 | mitochondrial fusion | 19 | 7 | 1.30E-03 |
| GO:0045088 | regulation of innate immune response | 300 | 21 | 1.30E-03 |
| GO:0034497 | protein localization to phagophore assembly site | 13 | 5 | 1.31E-03 |
| GO:0006610 | ribosomal protein import into nucleus | 8 | 4 | 1.31E-03 |
| GO:0075522 | IRES-dependent viral translational initiation | 10 | 4 | 3.51E-03 |
| GO:0016055 | Wnt signaling pathway | 289 | 34 | 3.68E-03 |

### Alternative splicing events and consequences

Isoform switches identified from the DIU analysis can be further analyzed to characterize specific splicing events and potential consequences^31^. An example isoform switch in Sestrin 3 (*SESN3*) is shown in Fig. 1a. The down-used and up-used isoforms in *SESN3* have distinct splicing patterns that could result in multiple potential consequences at the RNA and protein level.

**Figure 1.**
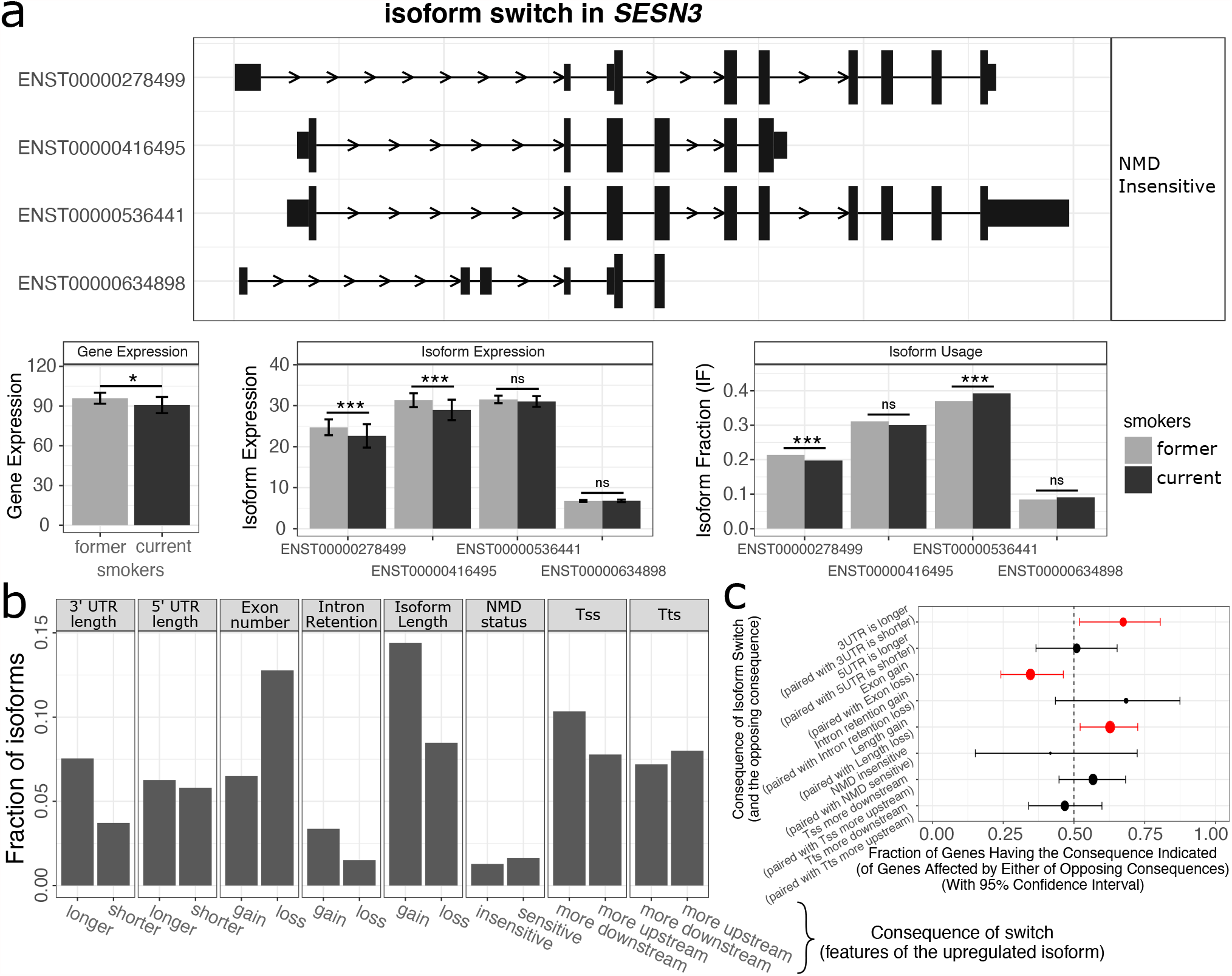
Smoking-associated isoform switches and consequences. An example of the identified isoform switches in the DIU analysis is shown in panel a, where only isoforms accounting for more than 5% of the gene expression are displayed. The statistical significance of DGE, DIE and DIU analysis is marked on the bar plots (*: q-value < 0.1, ***: q-value < 0.01, ns: nonsignificant). In panel b, pairwise comparisons between up-used and down-used isoforms for all tested genes are performed to assess specific consequences of isoform switches (e.g. 3′ UTR length is longer or shorter in up-used versus down-used isoforms from the same gene), and the fraction of DIU isoforms involved in a given type of switch consequence is shown. Panel c summarizes the net effects of these switch consequences at the gene level aggregated over all pairwise comparisons between up-used and down-used isoforms. Each gene will have a binary designation of its net switch consequence, and the fraction of genes with a particular designation and its confidence interval are shown. A binomial test is performed to assess the statistical significance of the gene fractions with respect to a null hypothesis of 0.5. The dot size is proportional to the number of genes whose DIU isoforms have a given type of switch consequences, and statistical significance of the binomial test is indicated by red colored dots. NMD: nonsense-mediated mRNA decay, identified from a premature termination codon >50nt upstream of the last exon-exon junction. Tss: transcription start site. Tts: transcription termination site.

By comparing splicing patterns between isoforms, we identified six categories of alternative splicing events prevalent in smoking-associated DIU isoforms, three of which (alternative transcription start site, alternative termination site, and intron retention) seem to be slightly more prevalent in the up-used isoforms (Supplementary Text 1). We next assessed the consequences of switching from down-used to up-used isoforms on eight isoform characteristics including UTR length, position of transcription start and termination site, intron retention, and sensitivity to NMD. We found isoform switching resulted in higher usage of isoforms that had longer overall length, longer 3′ UTRs, and fewer exons (p < 0.05 for all, Fig. 1b-c). In the example of *SESN3*, the up-used isoform has a longer isoform length due primarily to marked elongation of the 3′ UTR (7,742 nucleotides [nts] vs 107 nts in the down-used isoform).

### Smoking-associated increased usage of isoforms with extremely long 3′ UTRs

To further examine the significant isoform switch consequences related to length, we compared the length distribution of up-used, down-used, and non-DIU isoforms in genes identified through DIU analysis. We observed that isoforms up-used in current smokers were notably longer (median isoform lengths 2997 nts, 2323 nts, and 1221 nts for up-used, down-used, and non-DIU isoforms, respectively). The smoking-related transcript elongation occurred primarily in the coding region sequence (CDS) and 3′ UTRs but not in 5′ UTRs (Fig. 2). We also noted a strong correlation between CDS length and 3′ UTR length in all analyzed isoforms (Spearman rho = 0.78).

**Figure 2.**
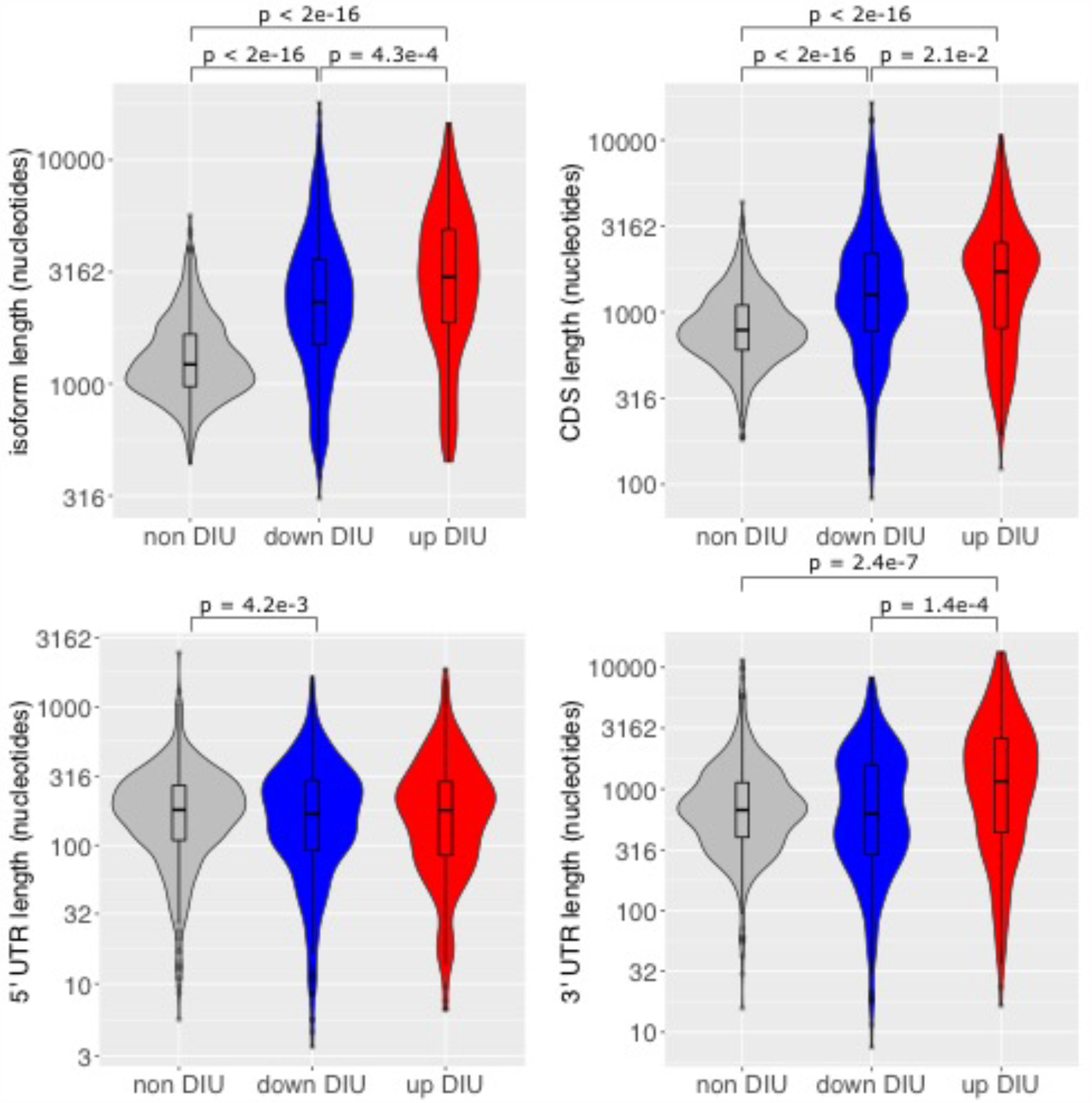
3′ UTR lengthening with up-used DIU isoforms. Isoforms were classified into three DIU categories (non-differentially used, down-used, up-used) according to their differential usage test statistics. Isoforms within each gene were grouped by category to compute average isoform length, 5′ UTR length, 3′ UTR length, and CDS length. These average lengths were compared across DIU categories using the Wilcoxon signed rank test, and the significant P-values are denoted in the violin plots. Significant differences in CDS and 3′ UTR length were observed, especially in up-used DIU isoforms.

Since these isoform-level analyses depend on the reliability of isoform expression estimation, we also performed differential exon usage (DEU) analysis on exonic part read counts directly supported by alignments. Exonic parts were derived from transcriptome annotations as described in^18^ and illustrated in Fig. 3a. We identified 126 up-used and 34 down-used exonic parts contained within 128 genes (Supplementary Table ST8). Forty-five percent (57/128) of these genes were also differentially expressed, 74% (42/57) of which were down-regulated.

**Figure 3.**
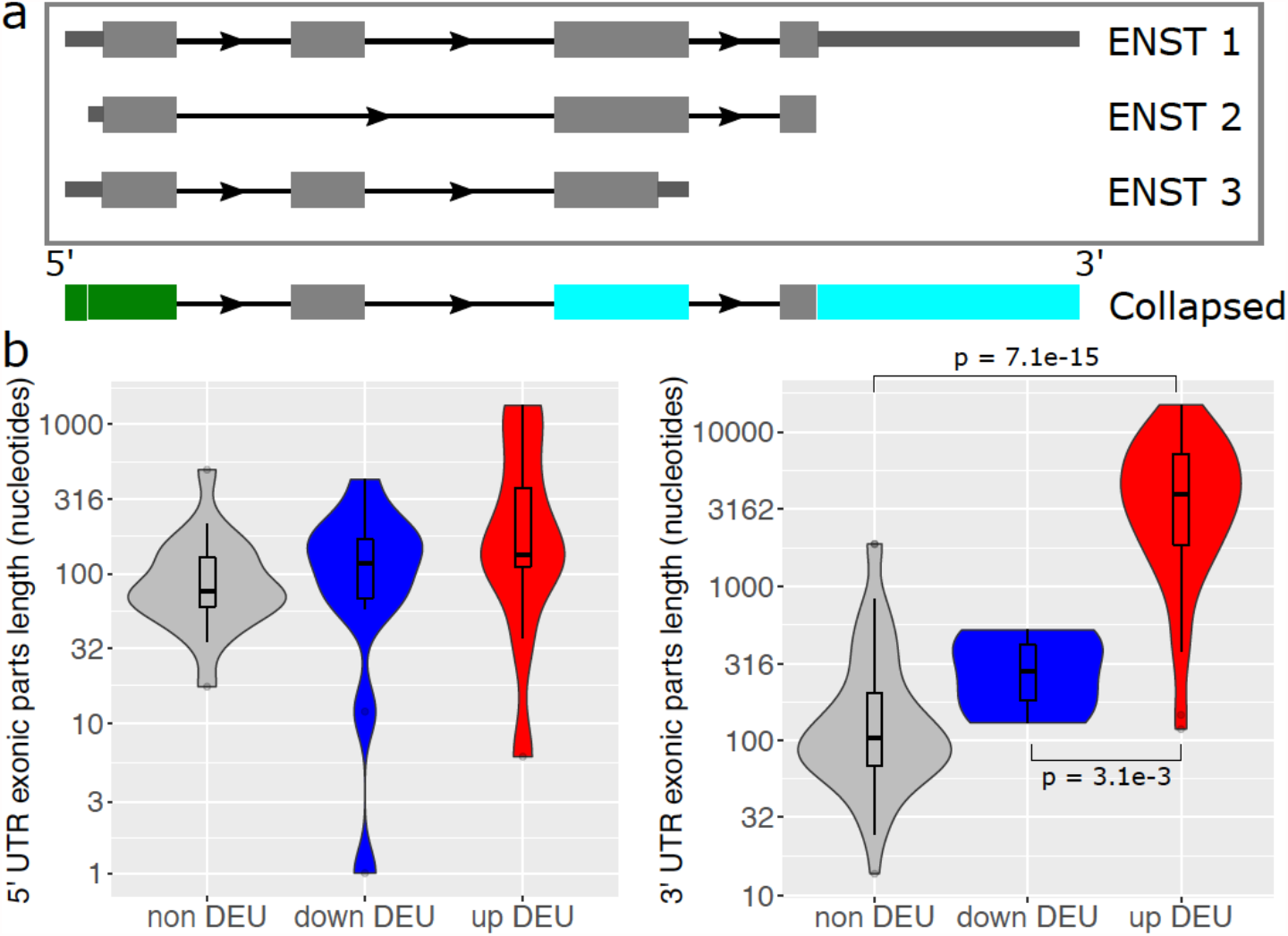
3′ UTR lengthening of up-used DEU exonic parts. Non-overlapping exonic parts were derived from collapsed Ensembl gene models, as illustrated in panel a. Exonic parts that overlap annotated 5′ and 3′ UTRs are colored in green and cyan, respectively. In panel b, exonic parts were classified into three DEU categories (non-differentially used, down-used, up-used) according to their differential usage test statistics. Exonic parts within each gene were grouped by category to compute the average 5′ UTR and 3′ UTR exonic parts length. These average lengths were compared between the three DEU categories using the Wilcoxon signed rank test, and the significant P-values are denoted in the violin plots. A significant increase in 3′ UTR exonic part length was observed in up-used DEUs.

Analysis on DEU exonic parts lengths confirmed the switch toward isoforms with extremely long 3′ UTRs (Fig. 3b). Differentially used 3′ UTRs (DEU 3′ UTRs) accounted for 40% (64/160) of all identified DEU exonic parts, nearly all (56/64) of which were up-used in current smokers. Of the genes containing a DEU 3′ UTR, about half (26/54) were differentially expressed with the large majority (19/26) showing decreased expression in current smokers. GO enrichment analysis of genes with up-used DEU 3′ UTRs identified over-representation of transcriptional regulation (e.g. polyadenylation and miRNA binding), Wnt-signaling and NF-kB signaling (Supplementary Table ST9). In summary, smoking results in marked 3′ UTR elongation that tends to be associated with a reduction in overall expression for the affected genes.

### Elongation of 3′ UTRs is not an artifact of transcript length bias

Transcript length bias in RNA-seq data analysis can arise when statistical power to detect differential expression is greater for longer isoforms, due to the fact that read counts are proportional to not only expression levels but also transcript lengths^38^. To determine whether the observed smoking-associated 3′ UTR elongation is driven by length bias, we compared our analysis on data where smoking status was randomly permuted. The results of this permutation analysis demonstrate that the magnitude of length-related effects observed in the non-permuted analysis far exceeds the effects seen with permutation, and that the directional preference of positive log-fold-changes for longer 3′ UTR isoforms in current smokers is absent in the permuted data (Fig. 4). These results indicate that the observed smoking-associated 3′ UTR elongation is not driven by transcript length bias.

**Figure 4.**
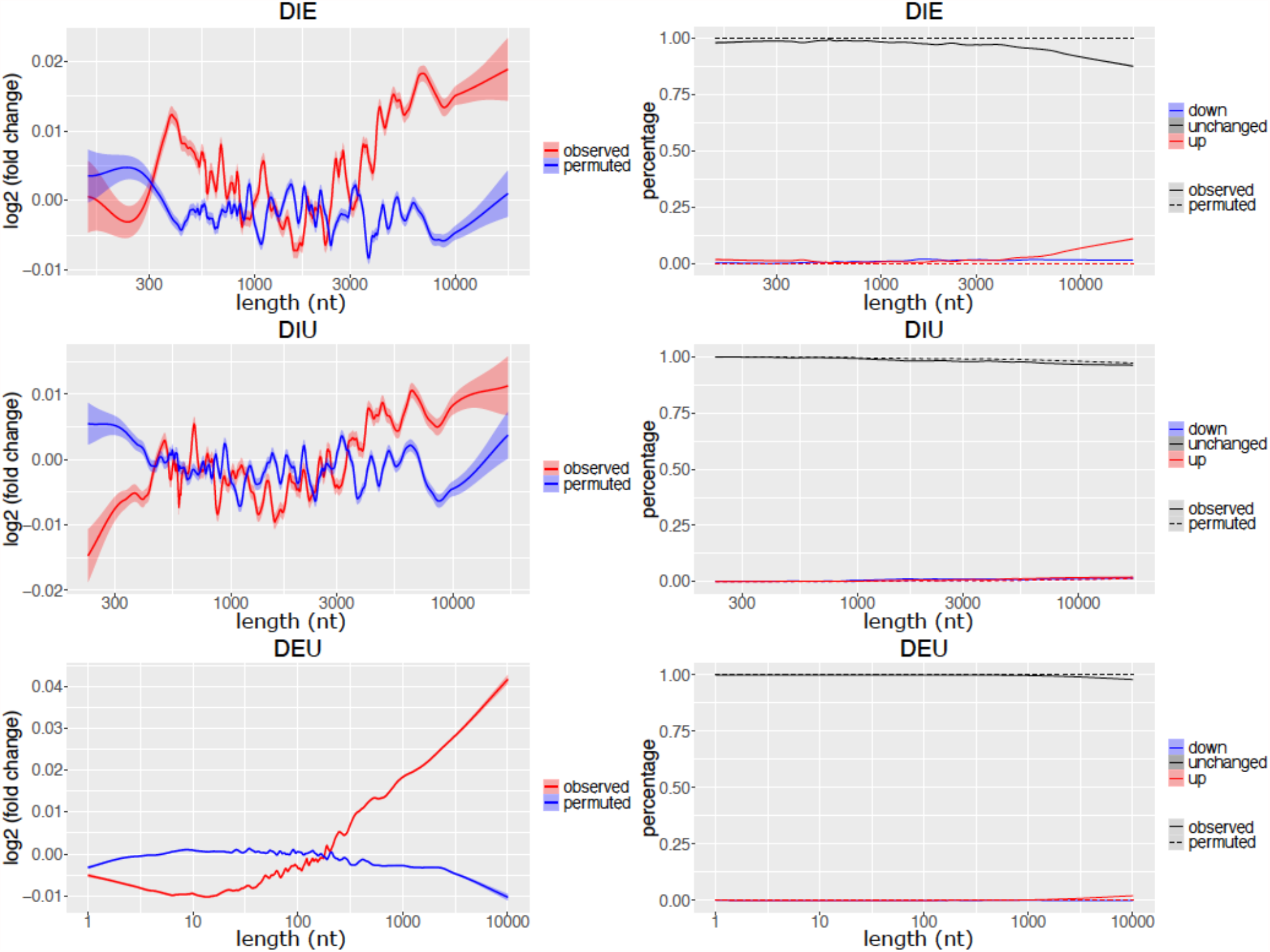
Directional preference in smoking-induced transcriptional regulation in observed versus permuted data. Left-hand panels demonstrate that a trend towards positive log fold changes (i.e. higher expression and usage in smokers) with longer features is present in the observed data but not the permuted data. Right-hand panels show the increasing percentage of features detected as up-regulated or up-used in smokers as the features become longer. Features were sorted by length, and statistics (average log fold changes and feature lengths) were computed from a sliding window of size 300 and step size 15 nts. A LOWESS curve was fit to these statistics and shown with 95% confidence interval in the left-hand panels for the three types of analysis (DIE, DIU and DEU) in both the observed and permuted data. In the right-hand panels, the percentage of features stratified by status of differential expression and usage were shown on the y axis.

### Alternative polyadenylation mediates 3′ UTR elongation

We next sought to determine whether smoking-associated 3′ UTR lengthening occurs in a controlled manner through transcriptional termination mechanisms involving alternative polyadenylation (APA). To test the hypothesis on alternative polyadenylation site (PAS) usage, we assessed whether annotated PAS are enriched within up-used 3′ UTRs. The majority of up-used 3′ UTRs (50/56) contained at least one annotated PAS, representing a thirtyfold enrichment over all other tested 3′ UTRs within the same genes (OR = 30.1, P-value < 0.001), and a twentyfold enrichment over 3′ UTRs across all genes. These enrichment scores remain highly significant when each 3′ UTR is trimmed to the last 100 nts at its 3′ end (Table 4, poly(A) sites). In contrast, PAS were identified in only 25% of down-used 3′ UTRs. We also observed at least one PAS in close proximity to the distal boundary of up-used 3′ UTRs (median distance of 7 nts), consistent with the hypothesis that the 3′ UTR extension is mediated through alternative usage of PAS.

**Table 4.** Summary of functional element enrichment in 3′ UTRs. Counts of each of the three types of functional elements in a given set of 3′ UTR exonic parts, as well as odds ratios and P-values of enrichment tests, are shown. The “full length” count method means counting the number of 3′ UTR exonic parts that harbor at least one functional element. The “last 100 bp” means counting similarly to “full length” except trimming all 3′ UTRs to include only the most distal 100 bp. The “last 100 bp density” counts the total number of functional elements in the last 100 bp of a given set of 3′ UTRs. The “within genes” and “across genes” denotes comparisons done entirely within genes containing up-used 3′ UTRs and comparisons done in all tested genes, respectively.

| Functional elements | Count method | Up-used 3' UTR |  | non-DEU 3' UTR |  |  |  |  |  |  |  |
| --- | --- | --- | --- | --- | --- | --- | --- | --- | --- | --- | --- |
|  |  |  |  | within genes |  |  |  | across genes |  |  |  |
|  |  | yes | no | yes | no | OR | P-value | yes | no | OR | P-value |
| poly(A) sites | full length | 50 | 6 | 117 | 425 | 30.08 | < 2.2e-16 | 23380 | 57386 | 20.45 | < 2.2e-16 |
|  | last 100 bp | 36 | 20 | 113 | 429 | 6.81 | 5.14E-11 | 20930 | 59836 | 5.15 | 1.88E-09 |
|  | last 100 bp density | 47 | 0 | 151 | 0 | 3.01 | 2.13E-09 | 26213 | 0 | 2.59 | 1.26E-08 |
| ARE | full length | 51 | 5 | 76 | 466 | 61.92 | < 2.2e-16 | 17854 | 62912 | 35.94 | < 2.2e-16 |
|  | last 100 bp | 24 | 32 | 58 | 484 | 6.23 | 1.13E-08 | 11731 | 69035 | 4.41 | 2.88E-07 |
|  | last 100 bp density | 35 | 0 | 70 | 0 | 4.84 | 1.12E-11 | 14262 | 0 | 3.54 | 4.67E-10 |
| miRNA binding sites | full length | 36 | 20 | 88 | 454 | 9.23 | 7.09E-14 | 15172 | 65594 | 7.78 | 1.03E-13 |
|  | last 100 bp | 13 | 43 | 83 | 459 | 1.67 | 1.28E-01 | 10769 | 69997 | 1.97 | 4.57E-02 |
|  | last 100 bp density | 33 | 0 | 164 | 0 | 1.95 | 1.27E-03 | 20823 | 0 | 2.29 | 2.78E-05 |

To ascertain whether there were any differences in strength of PAS in up-used 3′ UTRs relative to PAS in other 3′ UTRs in the same genes, we examined the frequency of the canonical poly(A) motif (AATAAA) as a surrogate for overall PAS strength. We focused on externally verified PAS within the last 100 nts of a 3′ UTR exonic part, and we counted instances in which AATAAA motifs were located within 60 nts upstream of a PAS. We found PASs in up-used 3′ UTRs had a higher frequency of AATAAA motifs than PAS in non-DEU 3′ UTRs from the same genes (44.7% versus 29.8%). The presence of AATAAA motifs was also correlated to exonic part differential usage P-values (Spearman rho = 0.19) and log-fold-changes (Spearman rho = 0.26). A similar pattern was observed for another strong poly(A) motif TTTTTTTTT (Fig. 5 a-c).

**Figure 5.**
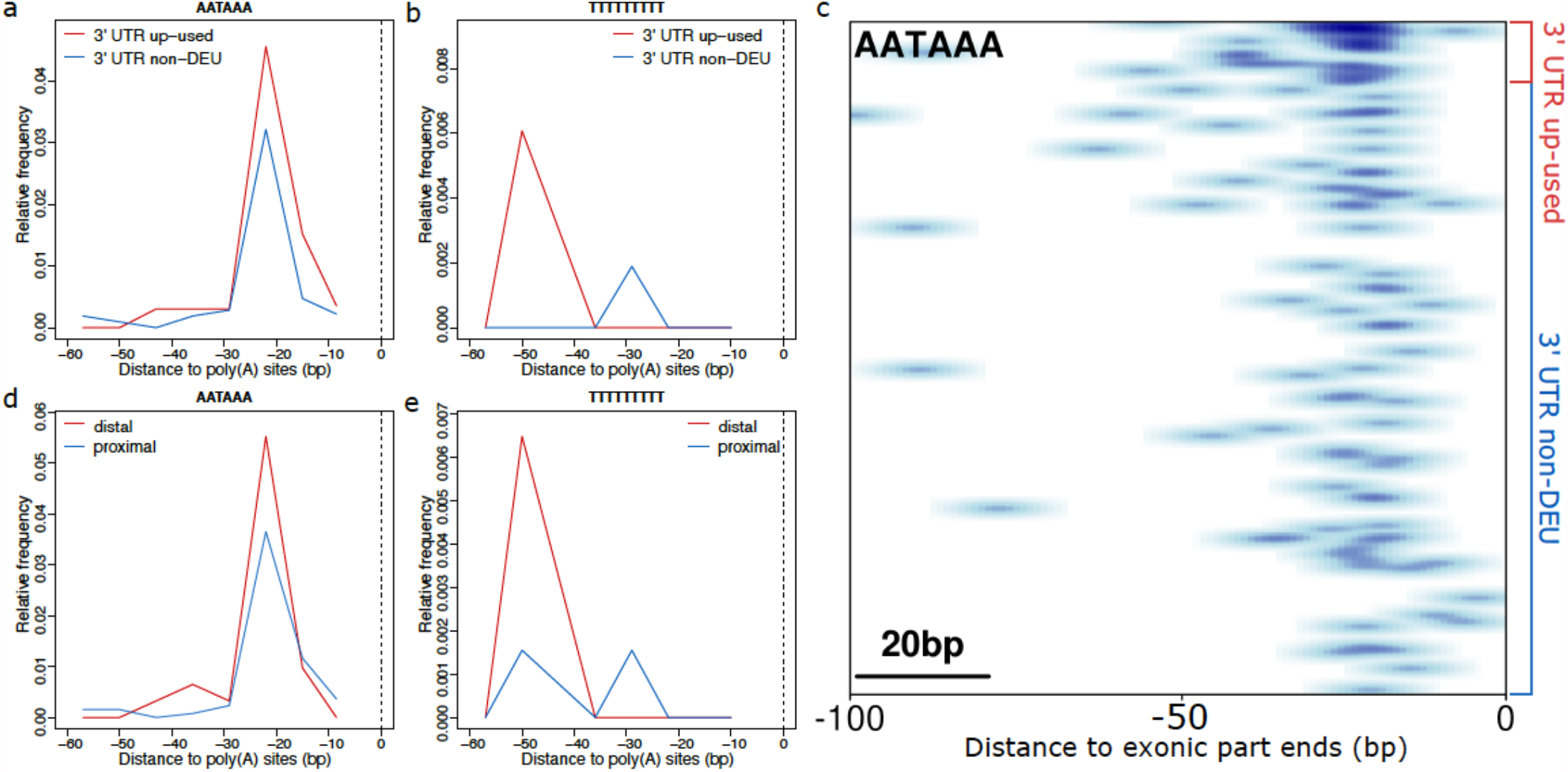
Positional frequency of poly(A) motifs upstream of polyadenylation sites (PASs). Genomic sequences upstream of PASs are extracted, and the frequencies of poly(A) motifs at each base position are computed and smoothed in the visualization. In panels a-b, PASs are categorized according to the DEU analysis of the 3′ UTRs harboring these sites. In panels d-e, PASs are categorized as distal and proximal depending on their location relative to the annotated end of the gene. In panels a-b and d-e, the dashed lines mark the position of experimentally determined PAS cleavage sites in 3′ UTR exonic parts from genes containing up-used 3′ UTRs. In panel c, each row represents the last 100 nucleotides of a 3′ UTR exonic part, ordered by DEU P-values.

To determine the localization of our DEU 3′ UTRs, we classified all exonic parts in genes containing DEU 3′ UTRs as distal (located at the gene end) and proximal (located at an upstream 3′ UTR). Fifty-two percent (29/56) of DEU 3′ UTRs were distal, and the frequency of the canonical poly(A) motif in these genes was highest in distal PASs (52.3%), compared to 35.9% for proximal PASs. Similarly, we found the highest frequency of poly(A) motif TTTTTTTTT at distal PASs. The positional frequencies of these motifs are shown in Fig. 5 d-e. This analysis highlights the predominant localization of DEU 3′ UTRs at gene ends with strong distal poly(A) signals.

### Enrichment of functional regulatory elements in smoking-elongated 3′ UTRs

3′ UTRs often harbor functional binding sites that regulate mRNA stability and localization, and previous work has shown that some transcripts with longer 3′ UTRs harbor repressive elements in extended 3′ UTR regions^39^. These functional sites often reside in adenylate-uridylate (AU)-rich elements that serve as regulatory hotspots characterized by joint binding of regulatory factors such as RBPs and miRNAs^40^.

Using the core pentamer motif of AREs (AUUUA), we found that AREs are significantly enriched in up-used 3′ UTRs relative to non-smoking associated 3′ UTRs (OR = 35.9, P-value < 0.001). When considering the density of AREs per unit length of 3′ UTR, ARE sites also occur at significantly higher frequency in up-used 3′ UTRs. We also observed enrichment of Targetscan predicted miRNA binding sites (OR = 7.8, P-value < 0.001) within up-used 3′ UTRs (Table 4). The chance of co-occurrence of these functional elements (including PAS) in up-used 3′ UTRs is significantly higher (Supplementary Table ST10). Positional frequency analysis clearly demonstrates an enriched distribution of PASs, AREs, and miRNA binding sites over the elongated 3′ UTRs, especially at the distal end (Fig. 6).

**Figure 6.**
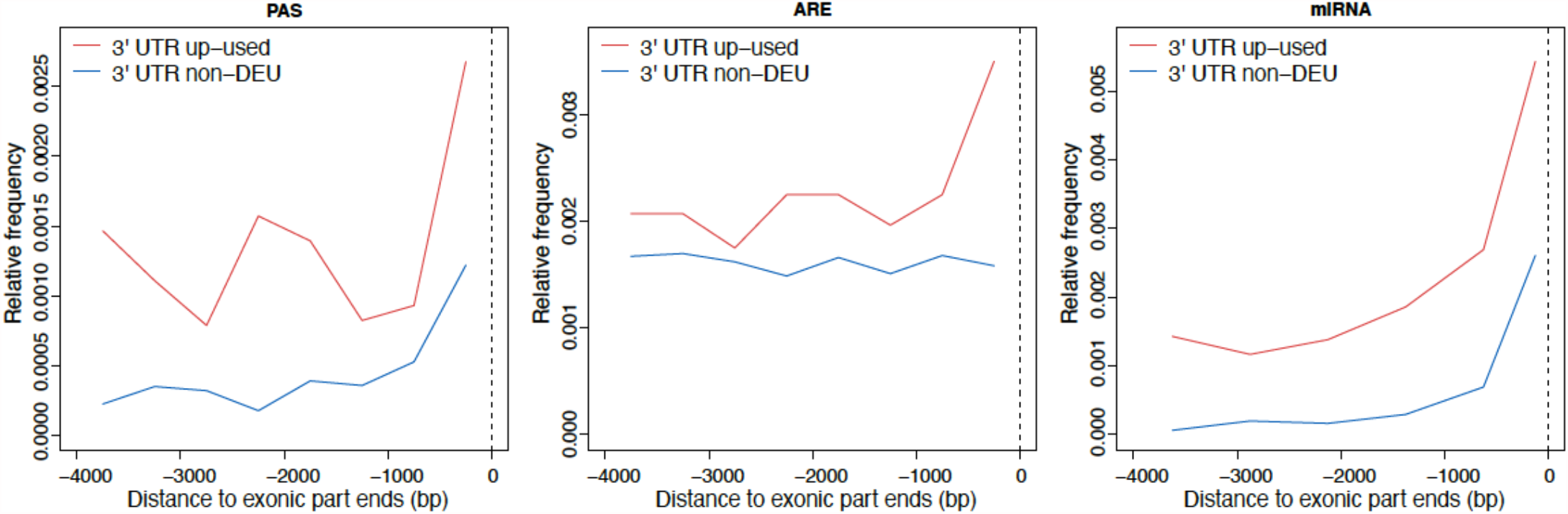
Positional frequency of functional elements in 3′ UTRs. Genomic sequences up to 4 kb upstream of the 3′ UTR exonic part ends are extracted, and the frequencies of functional elements (PAS, ARE, miRNA) at each base position are computed and smoothed in the visualization. The exonic parts analyzed here include all 3′ UTRs from genes containing up-used 3′ UTRs.

Extending the global enrichment analysis to individual regulatory factors, we identified five miRNAs and three RBPs whose binding sites were enriched in up-used 3′ UTRs. To explore putative coordination between these miRNAs and RBPs, a regulatory network of these entities and their target genes were constructed. Using a community detection algorithm, we identified five communities (modularity score 0.32) of dense connections, including four connected communities and one isolated RBP community (MATR3). Interestingly, *AGO2*, a member of the largest community, is a target for both the top 2 miRNA candidates and the top 2 RBP candidates, suggesting that these miRNAs and RBPs may act in a coordinated manner in post-transcriptional regulation of *AGO2* and other target genes (Fig. 7). AGO2 protein is essential to miRNA and siRNA-mediated post-transcriptional gene-silencing, and the most distal 3′ UTR of *AGO2* is up-used in response to smoking (q-value = 7.78e-8).

**Figure 7.**
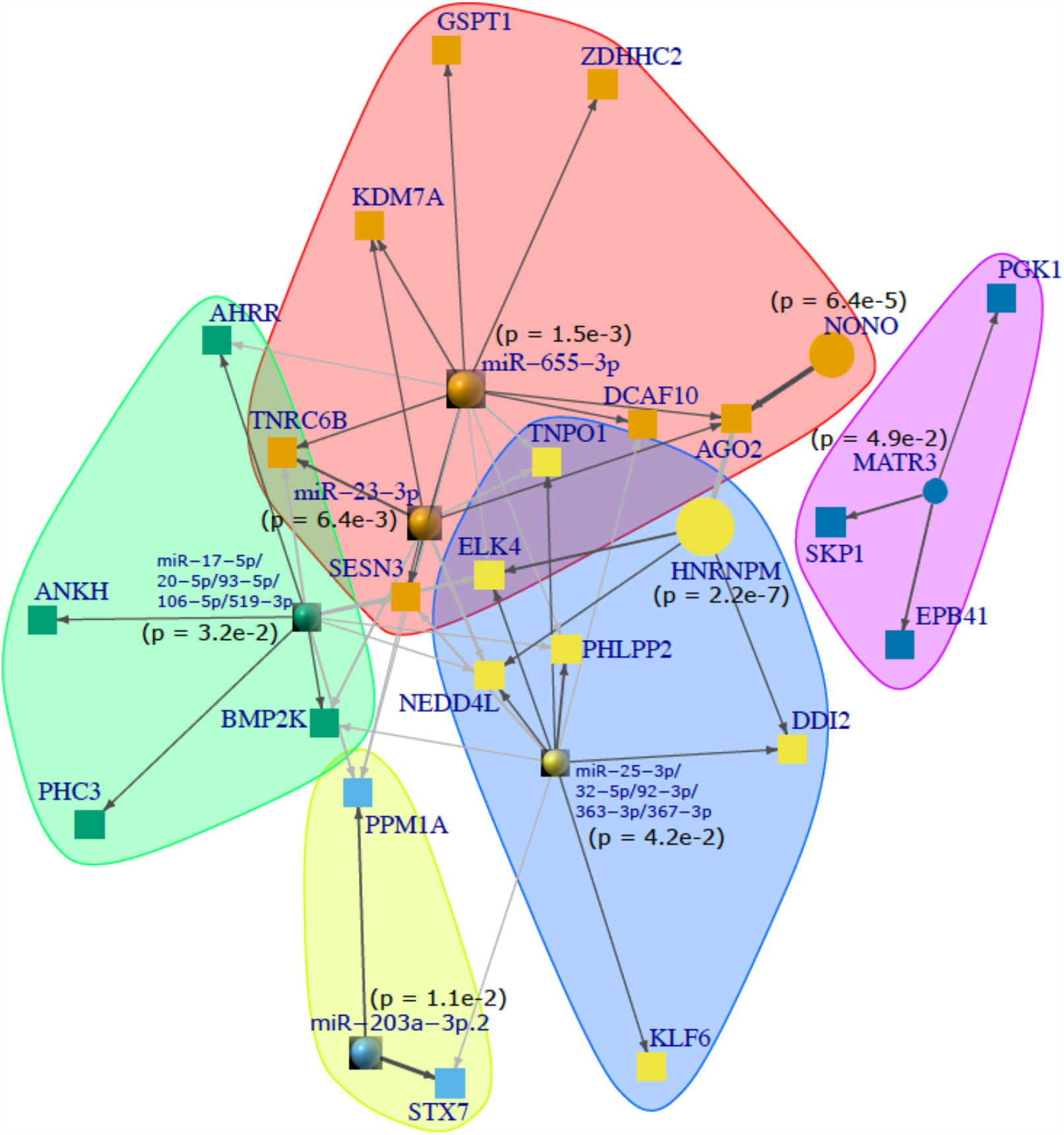
Regulatory network of micro-RNAs (miRNAs), RNA-binding proteins (RBPs) and their target genes with up-used 3′ UTRs. Candidate regulatory factors (5 miRNAs and 3 RBPs) were identified from enrichment tests of binding sites from TargetScan and e-CLIP experiments in up-used 3′ UTRs. Five network communities are designated by the node coloring and shaded polygons. p-values from the binomial enrichment tests are shown. Node size is proportional to the -log10 transformed binomial p-values. Node shape: sphere = miRNA; circle = RBP; square = target gene. Edge width is proportional to the number of binding sites. Edge color designates the within (black) or between (gray) community links.

### Alternative polyadenylation is implicated in smoking-related human diseases and traits

To relate smoking-induced alternative polyadenylation with human diseases and traits, we first performed eQTL analysis to identify genetic variants within a 1MB *cis* window associated with the expression level of smoking-related DEU 3′ UTRs. We found 2,840 significant QTLs at 5% FDR for 29 DEU 3′ UTRs in 25 genes. The majority (2582/2840 = 90.9%) of these QTLs were specifically associated with the expression level of 3′ UTR rather than the gene expression level. We then cross-referenced these QTLs against the NHGRI-EBI GWAS catalog^41,42^ and identified 79 GWAS variants that were significantly associated with expression levels of DEU 3′ UTRs in 11 genes. The most significant QTLs were associated with the up-used 3′ UTR in *ERAP1* (Supplementary Table ST11), an endoplasmic reticulum–expressed aminopeptidase that trims peptides for presentation by MHC class I molecules^43^. The minor allele of the lead QTL variant for *ERAP1*, rs7063, disrupts a canonical poly(A) motif AATAAA for the proximal poly(A) site, leading to increased usage of the distal poly(A) site and an isoform switch from the shorter isoform ENST00000443439 to the longer isoform ENST00000296754 with extended 3′ UTR (Fig. 8a-c). Although rs7063 is not cataloged in the NHGRI-EBI GWAS database, it has linkage disequilibrium (LD) to various degrees with nearby QTLs and GWAS variants including those associated with protein expression levels, alcohol dependence, ankylosing spondylitis and psoriasis (Fig. 8d). These results implicate alternative polyadenylation in posttranscriptional protein level modulation and smoking-related diseases and traits^44–46^.

**Figure 8.**
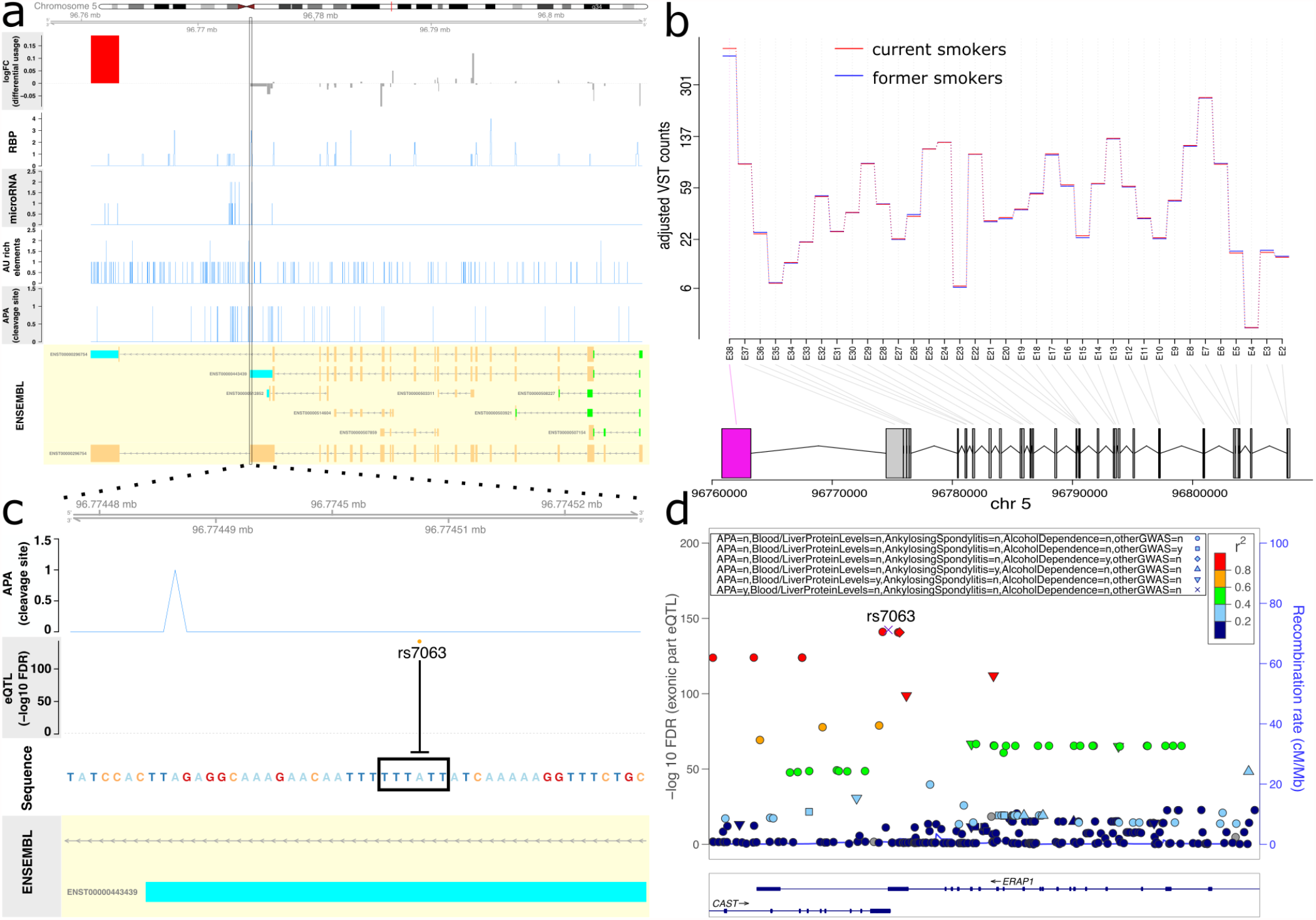
Genetic effects on alternative polyadenylation in *ERAP1*. Panel a shows sequentially in each row the differential usage log fold changes for the exonic parts, the coverage of RNA binding protein (RBP), miRNA, AU-rich elements (ARE), and alternative polyadenylation (APA) cleavage sites, and the Ensembl gene model for *ERAP1*. The exonic parts differential usage pattern is further illustrated in panel b using variance stabilized transformed (VST) counts adjusted for covariates. Panel c highlights the genetic variant directly disrupting the canonical poly(A) motif at the proximal poly(A) site. A LocusZoom plot is displayed in panel d, showing the eQTL FDR for the association of SNPs with the up-used 3′ UTR of *ERAP1*. The SNPs are colored according to linkage disequilibrium with the lead eQTL variant rs7063, and are annotated based on the effects on APA motifs and annotations in NHGRI-EBI GWAS catalog (n and y in the top legend means lacking and having effect/association, respectively).

## Discussion

Cigarette smoking increases susceptibility to many diseases including chronic obstructive pulmonary disease, cardiovascular disease, and multiple cancers. While the epidemiologic association of smoking to these disease risks is well-established, the underlying molecular basis is not fully understood, and the effects of smoking on alternative isoform regulation and posttranscriptional modulation have not been previously described. In a large cohort of current and former smokers, we used whole-blood RNA-seq to characterize the alternative splicing mechanisms and likely functional consequences of smoking-associated isoform switching. We demonstrated that smoking results in marked 3′ UTR elongation via alternative polyadenylation of genes enriched for specific biological pathways with disease implications. This 3′ UTR lengthening leads to the acquisition of post-transcriptional regulatory sites and is often associated with decreased overall expression of the affected genes.

The effect of smoking on gene expression in blood has been well-described^14,47–50^. The largest meta-analysis of smoking and blood transcriptome included 10,233 subjects, identifying 1,270 differentially expressed genes^50^. Our top associated genes were consistent with these previous studies. The only previous large-scale study related to alternative splicing in smoking was published on an earlier, smaller set of RNA-seq data from COPDGene^14^. This study identified 9 instances of DEU events but did not pursue analysis on isoform expression changes and switches, and the statistical power of that study was insufficient to systematically characterize alternative isoform regulation and posttranscriptional modulation in smoking. Expanding to twice as many subjects in the current study enabled us to identify hundreds of genes and biological pathways affected by smoking-associated isoform switching and APA events.

APA is a major RNA-processing mechanism that generates distinct 3ʹ termini on mRNAs and other RNA polymerase II transcripts, and contributes to human diseases including cancer, immunological and neurological diseases^51^. APA plays an important role in the cellular response to oxidative stress, heat shock and starvation^52^. Various kinds of environmental stress have been shown to increase utilization of distal polyadenylation sites^53^ and lead to transcriptional readthrough beyond annotated gene ends^54^. These observations suggest that APA may be a common posttranscriptional mechanism employed by mammalian cells when rapid modulations of RNA and protein levels are required in response to cellular stress. Smoking could be one of a larger class of exposures that elicits this posttranscriptional stress response, and additional studies of RNA-protein binding, RNA stability and trafficking are needed to elucidate its full spectrum of posttranscriptional modulations.

While previous genome-wide association studies (GWAS) have identified numerous genetic variants associated with smoking and smoking-related phenotypes^55–60^, functional interpretation of these variants remains challenging. Genetic variants could directly alter poly(A) motifs and RBP binding sites to modulate APA events, and several studies have been undertaken in recent years to systematically map novel apaQTLs and their disease etiologies^61–64^. Our preliminary 3′ UTR eQTL analysis in the current study suggests APA as a potential molecular phenotype to link genetic variants to smoking-related human diseases and traits. Further systematic apaQTL studies are needed to identify APA-related genetic-environment interactions conferring disease susceptibility.

The strengths of this study are the large sample size of RNA-seq data and the genome-wide assessment of alternative isoform regulation and posttranscriptional modulation in smoking. Our CBC quantifications do not capture variability of immune cell subpopulations, limiting our ability to localize these effects to specific cell types. Some of our results may reflect underlying changes in unmeasured cell type subpopulations. In future studies, the use of single cell data (scRNA-seq) or cell type deconvolution methods may provide additional insights. scRNA-seq may offer unique advantage in studying APA as the most popular scRNA-seq protocols specifically sequence the 3ʹ end of transcripts^65^.

In conclusion, our findings from 1,221 current and former smokers demonstrate widespread effects of smoking on alternative isoform regulation, highlighting specifically posttranscriptional mechanisms of APA and 3′ UTR lengthening. In the future, when longitudinal follow-up data are available for these subjects, we may be able to relate these posttranscriptional events to prospective health outcomes, and develop APA biomarkers and therapeutic targets for smoking-related diseases^66^.

## Supporting information

Supplementary Methods for VST method

## Data Availability

Data have been deposited in GEO.

## Funding/Acknowledgements

This work was funded by R01 HL124233, R01 HL147326, R01 HL111527, U01 HL089897, U01 HL089856, R01HL125583, R01HL130512, R01 GM101237, R01 HL11152, K25HL140186 and K08HL141601. Research reported in this publication was supported by the NHLBI, NIGMS and FDA Center for Tobacco Products (CTP). The content is solely the responsibility of the authors and does not necessarily represent the official views of the NIH or the Food and Drug Administration.

## COPDGene® Investigators – Core Units

Administrative Center: James D. Crapo, MD (PI); Edwin K. Silverman, MD, PhD (PI); Barry J. Make, MD; Elizabeth A. Regan, MD, PhD

Genetic Analysis Center: Terri Beaty, PhD; Ferdouse Begum, PhD; Peter J. Castaldi, MD, MSc; Michael Cho, MD; Dawn L. DeMeo, MD, MPH; Adel R. Boueiz, MD; Marilyn G. Foreman, MD, MS; Eitan Halper-Stromberg; Lystra P. Hayden, MD, MMSc; Craig P. Hersh, MD, MPH; Jacqueline Hetmanski, MS, MPH; Brian D. Hobbs, MD; John E. Hokanson, MPH, PhD; Nan Laird, PhD; Christoph Lange, PhD; Sharon M. Lutz, PhD; Merry-Lynn McDonald, PhD; Dandi Qiao, PhD; Elizabeth A. Regan, MD, PhD; Edwin K. Silverman, MD, PhD; Emily S. Wan, MD; Sungho Won, PhD

Imaging Center: Juan Pablo Centeno; Jean-Paul Charbonnier, PhD; Harvey O. Coxson, PhD; Craig J. Galban, PhD; MeiLan K. Han, MD, MS; Eric A. Hoffman, Stephen Humphries, PhD; Francine L. Jacobson, MD, MPH; Philip F. Judy, PhD; Ella A. Kazerooni, MD; Alex Kluiber; David A. Lynch, MB; Pietro Nardelli, PhD; John D. Newell, Jr., MD; Aleena Notary; Andrea Oh, MD; Elizabeth A. Regan, MD, PhD; James C. Ross, PhD; Raul San Jose Estepar, PhD; Joyce Schroeder, MD; Jered Sieren; Berend C. Stoel, PhD; Juerg Tschirren, PhD; Edwin Van Beek, MD, PhD; Bram van Ginneken, PhD; Eva van Rikxoort, PhD; Gonzalo Vegas Sanchez-Ferrero, PhD; Lucas Veitel; George R. Washko, MD; Carla G. Wilson, MS;

PFT QA Center, Salt Lake City, UT: Robert Jensen, PhD

Data Coordinating Center and Biostatistics, National Jewish Health, Denver, CO: Douglas Everett, PhD; Jim Crooks, PhD; Katherine Pratte, PhD; Matt Strand, PhD; Carla G. Wilson, MS

Epidemiology Core, University of Colorado Anschutz Medical Campus, Aurora, CO: John E. Hokanson, MPH, PhD; Gregory Kinney, MPH, PhD; Sharon M. Lutz, PhD; Kendra A. Young, PhD

Mortality Adjudication Core: Surya P. Bhatt, MD; Jessica Bon, MD; Alejandro A. Diaz, MD, MPH; MeiLan K. Han, MD, MS; Barry Make, MD; Susan Murray, ScD; Elizabeth Regan, MD; Xavier Soler, MD; Carla G. Wilson, MS

Biomarker Core: Russell P. Bowler, MD, PhD; Katerina Kechris, PhD; Farnoush Banaei-Kashani, Ph.D COPDGene® Investigators – Clinical Centers Ann Arbor VA: Jeffrey L. Curtis, MD; Perry G. Pernicano, MD

Baylor College of Medicine, Houston, TX: Nicola Hanania, MD, MS; Mustafa Atik, MD; Aladin Boriek, PhD; Kalpatha Guntupalli, MD; Elizabeth Guy, MD; Amit Parulekar, MD

Brigham and Women’s Hospital, Boston, MA: Dawn L. DeMeo, MD, MPH; Alejandro A. Diaz, MD, MPH; Lystra P. Hayden, MD; Brian D. Hobbs, MD; Craig Hersh, MD, MPH; Francine L. Jacobson, MD, MPH; George Washko, MD

Columbia University, New York, NY: R. Graham Barr, MD, DrPH; John Austin, MD; Belinda D’Souza, MD; Byron Thomashow, MD

Duke University Medical Center, Durham, NC: Neil MacIntyre, Jr., MD; H. Page McAdams, MD; Lacey Washington, MD

Grady Memorial Hospital, Atlanta, GA: Eric Flenaugh, MD; Silanth Terpenning, MD

HealthPartners Research Institute, Minneapolis, MN: Charlene McEvoy, MD, MPH; Joseph Tashjian, MD

Johns Hopkins University, Baltimore, MD: Robert Wise, MD; Robert Brown, MD; Nadia N. Hansel, MD, MPH; Karen Horton, MD; Allison Lambert, MD, MHS; Nirupama Putcha, MD, MHS

Lundquist Institute for Biomedical Innovationat Harbor UCLA Medical Center, Torrance, CA: Richard Casaburi, PhD, MD; Alessandra Adami, PhD; Matthew Budoff, MD; Hans Fischer, MD; Janos Porszasz, MD, PhD; Harry Rossiter, PhD; William Stringer, MD

Michael E. DeBakey VAMC, Houston, TX: Amir Sharafkhaneh, MD, PhD; Charlie Lan, DO

Minneapolis VA: Christine Wendt, MD; Brian Bell, MD; Ken M. Kunisaki, MD, MS

National Jewish Health, Denver, CO: Russell Bowler, MD, PhD; David A. Lynch, MB

Reliant Medical Group, Worcester, MA: Richard Rosiello, MD; David Pace, MD

Temple University, Philadelphia, PA: Gerard Criner, MD; David Ciccolella, MD; Francis Cordova, MD; Chandra Dass, MD; Gilbert D’Alonzo, DO; Parag Desai, MD; Michael Jacobs, PharmD; Steven Kelsen, MD, PhD; Victor Kim, MD; A. James Mamary, MD; Nathaniel Marchetti, DO; Aditi Satti, MD; Kartik Shenoy, MD; Robert M. Steiner, MD; Alex Swift, MD; Irene Swift, MD; Maria Elena Vega-Sanchez, MD

University of Alabama, Birmingham, AL: Mark Dransfield, MD; William Bailey, MD; Surya P. Bhatt, MD; Anand Iyer, MD; Hrudaya Nath, MD; J. Michael Wells, MD

University of California, San Diego, CA: Douglas Conrad, MD; Xavier Soler, MD, PhD; Andrew Yen, MD

University of Iowa, Iowa City, IA: Alejandro P. Comellas, MD; Karin F. Hoth, PhD; John Newell, Jr., MD; Brad Thompson, MD

University of Michigan, Ann Arbor, MI: MeiLan K. Han, MD MS; Ella Kazerooni, MD MS; Wassim Labaki, MD MS; Craig Galban, PhD; Dharshan Vummidi, MD

University of Minnesota, Minneapolis, MN: Joanne Billings, MD; Abbie Begnaud, MD; Tadashi Allen, MD

University of Pittsburgh, Pittsburgh, PA: Frank Sciurba, MD; Jessica Bon, MD; Divay Chandra, MD, MSc; Carl Fuhrman, MD; Joel Weissfeld, MD, MPH

University of Texas Health, San Antonio, San Antonio, TX: Antonio Anzueto, MD; Sandra Adams, MD; Diego Maselli-Caceres, MD; Mario E. Ruiz, MD; Harjinder Singh

