## Supplementary Text for "Cigarette Smoking-Associated Isoform Switching and 3’ UTR Lengthening Via Alternative Polyadenylation": SupplementaryText1_currentSmokingUTR3_05142021.docx

^1^Channing Division of Network Medicine, Brigham and Women’s Hospital, Boston, MA, USA; ^2^Harvard Medical School, Boston, MA, USA; ^3^Division of Pulmonary and Critical Care Medicine, Brigham and Women’s Hospital, Boston, MA, USA; ^4^Pulmonary Drug Discovery Laboratory, Bayer US LLC. Pharmaceuticals, Research & Development, Boston, MA, USA; ^5^Northeastern University, Boston, MA, USA; ^6^Division of Pulmonary and Critical Care Medicine, National Jewish Health, Denver, CO, USA; ^7^Department of Biology, University of North Carolina at Chapel Hill, Chapel Hill, NC, USA; ^8^Division of General Internal Medicine and Primary Care, Brigham and Women’s Hospital, Boston, MA, USA

**Supplemental Methods**

**Total RNA extraction**

Total RNA was extracted from PAXgene TM Blood RNA tubes using the Qiagen PreAnalytiX PAXgene Blood miRNA Kit (Qiagen, Valencia, CA). The extraction protocol was performed either manually or with the Qiagen QIAcube extraction robot. Extracted RNA samples with RNA Integrity number (RIN) > 6 and concentration > =25 μg/ul were sequenced. The median RIN of sequenced samples was 8.3.

**cDNA library construction and sequencing**

Globin reduction and cDNA library preparation for total RNA was performed with the Illumina TruSeq Stranded Total RNA with Ribo-Zero Globin kit (Illumina, Inc., San Diego, CA). Library quality control included quantification with picogreen, size analysis on an Agilent Bioanalyzer or Tapestation 2200 (Agilent, Santa Clara, CA), and qPCR quantitation against a standard curve. 75 bp paired end reads were generated on Illumina sequencers. Samples were sequenced to an average depth of 20 million reads.

**Sequencing read alignment and quality control**

Reads were trimmed of TruSeq adapters using Skewer with default parameters^1^. Trimmed reads were aligned to the GRCh38 genome using the STAR aligner^2^. Quality control was performed using the FastQC^3^ and RNA-SeQC programs^4^. Samples were included for subsequent analysis if they had >10 million total reads, >80% of reads mapped to the reference genome, XIST and Y chromosome expression were consistent with reported gender, <10% of R1 reads were the sense orientation, Pearson correlation was > = 0.9 with samples in the same library construction batch, and genotype calls between variants called from RNA sequencing reads and DNA genotyping were concordant.

**Filtering and normalization**

Genomic features (genes, isoforms or exonic parts) of low expression (average counts per million (CPM) < 0.2 or the number of subjects with CPM > 0.5 less than 50) or outlying expression (the number of subjects with CPM > 50,000 is larger than 0 but less than 50) were dropped from further analysis. Differences in sequencing depth and RNA library composition between subjects were normalized using the TMM^5^ procedure from the Bioconductor edgeR^6^ package (v3.24.3).

**Annotation of APA, AU-rich elements, miRNA and RBP binding sites**

The genomic locations of polyadenylation cleavage sites (PAS) derived from a total of 475,703,248 reads from 78 3’-end sequencing libraries generated with 7 different 3’ end sequencing protocols^7^ were obtained from <http://polyasite.unibas.ch> (version r1.0 archived at <http://polyasite-v1.scicore.unibas.ch/>). The genomic locations of the core pentamer motif AUUUA of the AU-rich elements (AREs) were obtained from the AREsite2 database (<http://nibiru.tbi.univie.ac.at/AREsite2/welcome>). Predicted conserved genomic binding sites of conserved miRNA families were downloaded from TargetScan release 7.2 (http://www.targetscan.org/vert_72/vert_72_data_download/Predicted_Target_Locations.default_predictions.hg19.bed.zip). Both PAS and miRNA annotations were converted from hg19 to hg38 genomic coordinates via the deepblue_liftover function from DeepBlueR package (v1.8.0).

e-CLIP data on RNA-protein binding sites were obtained via the ENCODE data portal (<https://www.encodeproject.org/search/?type=Experiment&status=released>) for 122 RBPs on Nov. 18, 2019. The download was restricted only to RBPs whose ``Target Category" in ENCODE matched ``RNA binding protein" and included assays from only K562 and HepG2 cell lines. Using the narrowPeak bed files with ``1, 2" listed in the biological replicates column in the downloaded ENCODE metadata file, we mapped binding peaks to transcript and gene ids as annotated in the Ensembl GTF.

**Supplemental Results**

**Serum cotinine level – boxplot**

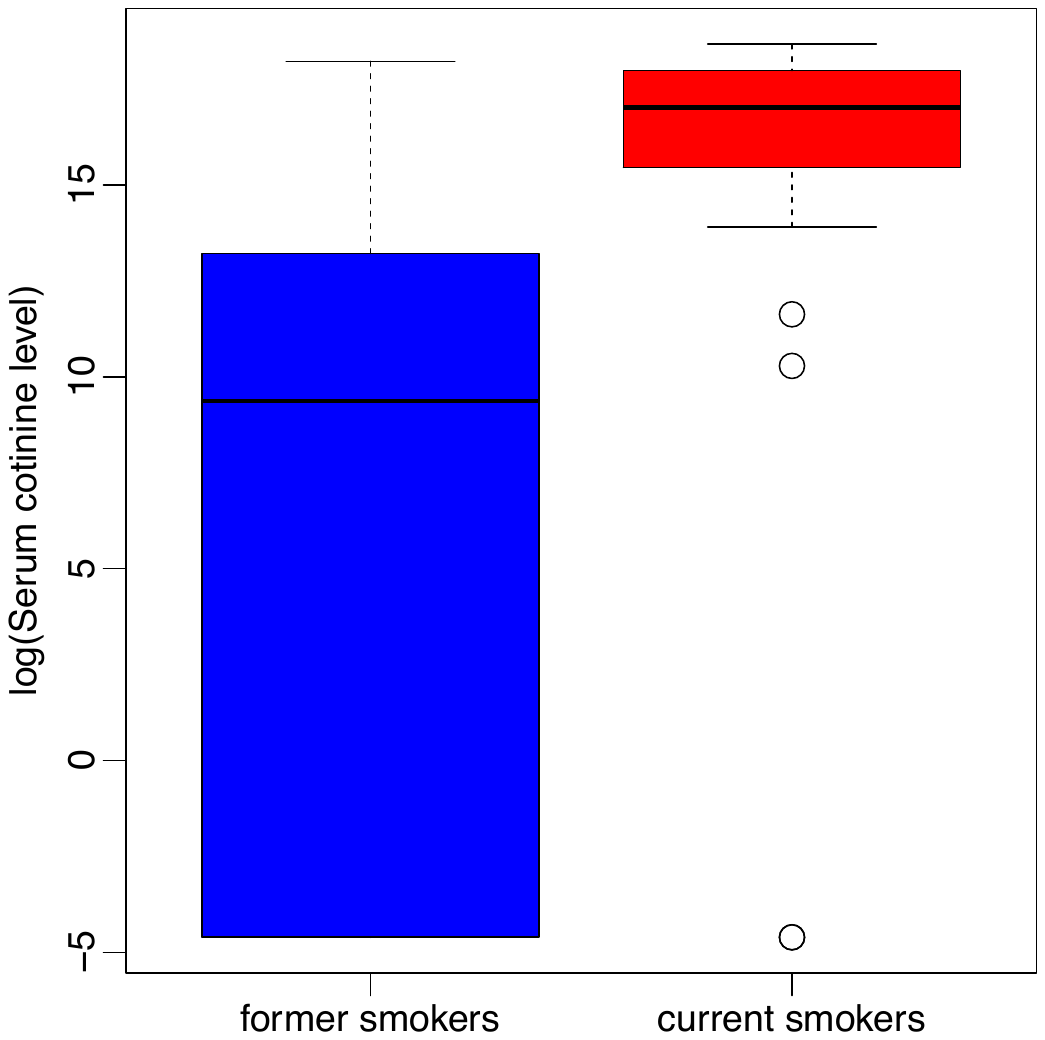

Fig. SF1. Validation of smoking status via serum cotinine measurements. In a subset of subjects (158 former and 46 current smokers included in the current study), measurements of serum cotinine levels confirmed the general accuracy of subjects’ self-reported smoking behavior in the COPDGene Study.

**Differential gene and isoform expression analysis – MA-plot**

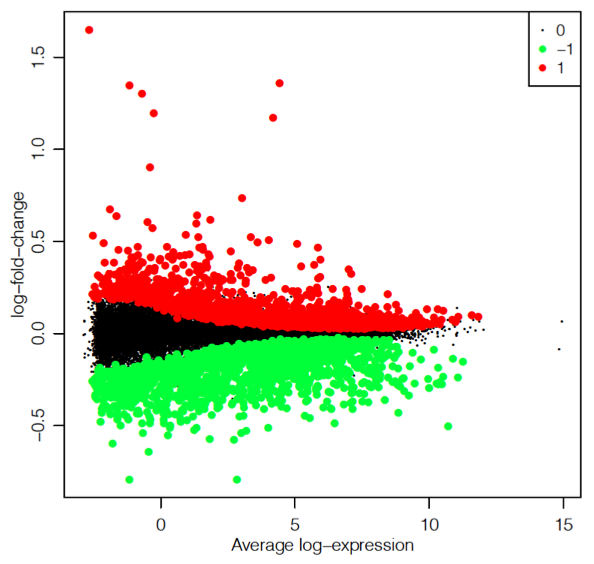

Fig. SF2. MA-plot for differential gene expression analysis. Each dot represents a gene, and red and green color denote genes significantly up-regulated and down-regulated at 10% FDR, respectively.

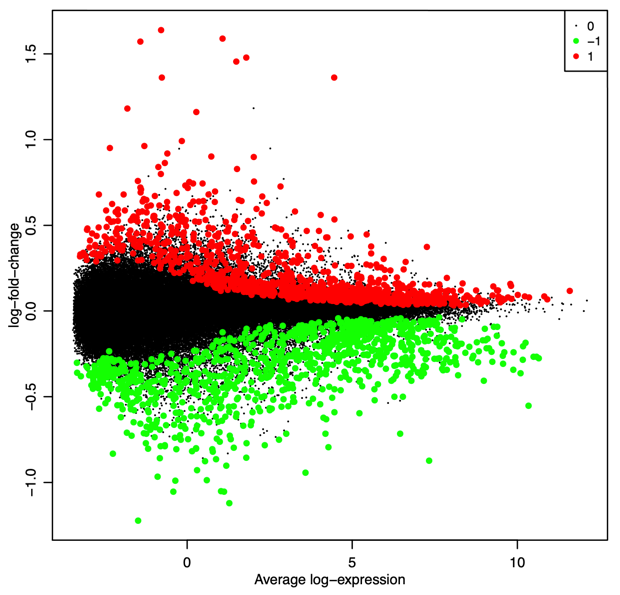

Fig. SF3. MA-plot for differential isoform expression analysis. Each dot represents a transcript, and red and green color denote transcripts significantly up-regulated and down-regulated at 10% FDR, respectively.

**Differential expression and usage analysis Venn diagram**

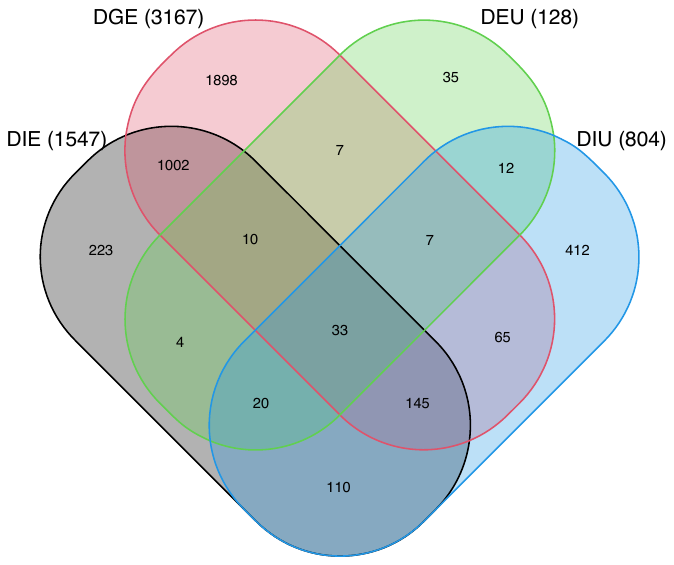

Fig. SF4. Venn diagram of genes identified from various types of differential expression and usage analysis. Numbers in parenthesis show marginal total number of genes from each type of analysis. DGE: differential gene expression; DIE: differential isoform expression; DIU: differential isoform usage; DEU: differential exon usage.

**Smoking-associated alternative splicing events**

As illustrated in Fig. SF5a, we can categorize alternative splicing events into ES (exon skipping), MES (multiple exon skipping), MEE (mutually exclusive exons), IR (intron retention), A5 (alternative 5’ splice site), A3 (alternative 3’ splice site), ATSS (alternative transcription start site), and ATTS (alternative transcription termination site)^8^. We found that nearly all of these splicing events are prevalent (> 45%) in smoking associated DIU isoforms, with the exception of IR and MEE (Fig. SF5b). Although splicing events of ATSS, ATTS and IR seem to be more prevalent in the up-used isoforms compared to the down-used ones, the differences were modest and not statistically significant when aggregated at gene level (Fig. SF5c).

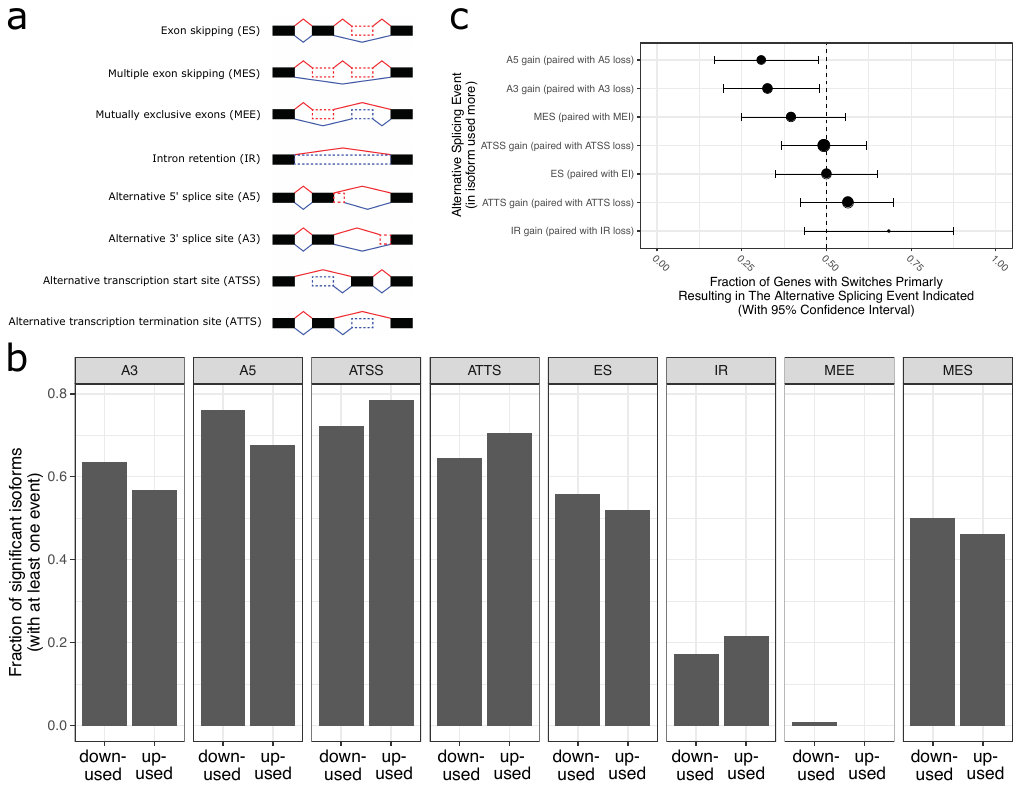

Fig. SF5. Smoking-associated isoform switches and alternative splicing. Categorization of alternative splicing events involved in two isoforms is illustrated in panel a (adapted from Vitting-Seerup et al.^9^). Panel b shows the fraction of DIU isoforms having a particular type of alternative splicing event stratified by the direction of isoform usage, where up-used and down-used indicate isoforms that are used more or less in smokers, respectively. Panel c summarizes the net gain or loss of these splicing events at gene level aggregated over pair-wise comparisons between up-used and down-used isoforms. Each gene will have a binary designation of net gain or loss, and the fraction of genes with a particular designation and its confidence interval are shown. A binomial test is performed to assess the statistical significance of the gene fractions with respect to a null hypothesis of 0.5. The dot size is proportional to the number of genes whose DIU isoforms have a given category of splicing events, and statistical significance of the binomial test is indicated by red colored dots.
