## Supplementary Methods for VST method for "Cigarette Smoking-Associated Isoform Switching and 3’ UTR Lengthening Via Alternative Polyadenylation": SupplementaryText2_VSTvoom_currentSmokingUTR3.pdf

### Variance Stabilizing Transformation with Voom

*Zhonghui Xu,*
*Peter Castaldi,*

*May 05, 2021*

#### Introduction

A number of statistical methods and tools have been developed for RNA-seq differential expression analysis. The limma-voom method [1] is a popular one that accounts for mean-variance relationship by estimating precision weights that can be entered into the linear modeling framework in the limma empirical Bayes pipeline. The linear modeling stratagies dramatically reduce the computational time, enabling limma-voom to process large number of RNA-seq samples that may otherwise be infeasible for competing methods modeling on negative binomial distribution.

RNA-seq data are known to display heteroscedasticity, and packages such as DESeq2 [2] and DEXSeq [3] offer **variance stabilizing transformation (VST)** to transform the count data to achieve approximate homoscedasticity. Such transformed RNA-seq data can be useful for visualization and machine learning applications. While the *voom* function in the limma package estimates a mean-variance trend, it operates primarily on the unit of log-cpm values and fits a nonparametric LOWESS curve to the trend. VST on counts under limma settings are not readily available. Here we describe an approximate but accurate procedure to derive dispersion estimates and VST from the limma-voom pipeline. Such procedure also unlocks the possibility to speed up negative binomial based modeling methods for large RNA-seq studies (e.g. replacing the *estimateDispersionsGeneEst* step in DESeq2 with the proposed procedure).

#### Method

For convenience, we use the same notations as in the voom paper [1]. Let $r_{gi}$ indicates read count for RNA samples $i = 1, 2, \cdots, n$, and genes $g = 1, 2, \cdots, G$. The library size of sample $i$ is the total number of mapped reads:

$$R_i = \sum_{g=1}^{G} r_{gi}$$

The log-counts per million (log-cpm) value is defined as:

$$y_{gi} = log_2\left(\frac{r_{gi} + 0.5}{R_i + 1} \times 10^6\right)$$

Let $\lambda_{gi} = E(r_{gi})$ be the expected read count under given experimental conditions. We assume the variance of the read count is related to the mean in a quadratic form as in a negative binomial distribution:

$$var(r_{gi}) = \lambda_{gi} + \phi_g \lambda_{gi}^2$$

Using Taylor's theorem, we can write the variance of the log-cpm value approximately as:

$$var(y_{gi}) \approx \frac{1}{(log(2))^2}\left(\frac{1}{\lambda_{gi}} + \phi_g\right)$$

The *voom* function in limma package estimates a gene-wise mean-variance relationship, and returns for each gene the average log-count and the square root of residual standard deviation from a linear model fitted on the normalized log-cpm values. We denote these two quantities as $\hat{\lambda}_g$ and $\hat{\sigma}_g$, respectively. We can derive gene-wise dispersion estimates as follows:

$$\hat{\phi}_g = (log(2))^2 \times \hat{\sigma}_g^4 - \frac{1}{2^{\hat{\lambda}_g}}$$

To model the dependence of dispersion on mean, we follow the parameterization in DEXSeq paper:

$$\phi_g = \frac{a_1}{2^{\lambda_g}} + a_0$$

To estimate the coefficients $a_0$ and $a_1$, we fit a gamma-family GLM model to the $\hat{\phi}_g$ and $\frac{1}{2^{\hat{\lambda}_g}}$ values using the function *parametricDispersionFit* available in the DESeq2 package. We can then write out the expanded form of the mean-variance function as:

$$var(r_{gi}) = \lambda_{gi} + (\frac{\hat{a}_1}{\lambda_{gi}} + \hat{a}_0)\lambda_{gi}^2 = \hat{a}_0\lambda_{gi}^2 + (1 + \hat{a}_1)\lambda_{gi}$$

A variance stabilizing transformation (VST) on the read count can thus be obtained by solving:

$$vst(r) = \int^r \frac{1}{\sqrt{\hat{a}_0\lambda^2 + (1 + \hat{a}_1)\lambda}} d_\lambda$$

By change of variables and knowing the derivative of inverse hyperbolic cosine function $\frac{d}{d_x} arccoshx = \frac{1}{\sqrt{x^2-1}}$, we can derive a VST solution as:

$$vst(r) = \frac{2}{\sqrt{\hat{a}_0}} \log(\sqrt{\hat{a}_0 r} + \sqrt{\hat{a}_0 r + \hat{a}_1 + 1})$$

This is equivalent to the VST function derived in the DEXSeq paper up to a constant. From the above VST function, it is clear that this transformation is essentially determined by the quantity $\frac{\hat{a}_1+1}{\hat{a}_0}$ up to a scaling and constant. In certain settings, it would be convenient to be able to transform back to the orginal read counts. So we further derive the inverse of the VST function as:

$$vst^{-1}(z) = \frac{1}{4\hat{a}_0}\left(e^{\frac{\sqrt{\hat{a}_0}}{2}z} - (\hat{a}_1 + 1)e^{-\frac{\sqrt{\hat{a}_0}}{2}z}\right)^2$$

An R package **voomst** has been created to implement the described method.

### Simulation

We simulate an RNA-seq data set with 5000 genes and 12 samples in two conditions. The true values for the parameters in the mean-dispersion relationship are set as: $a_0 = 0.1$ and $a_1 = 4$. We will demonstrate how well the DESeq2 negative binomial pipeline and the proposed limma-voom based pipeline estimate these dispersions and parameters. As we will see, these methods already perform well under this moderate sequencing depth and small sample size.

```
library(voomst)
library(DESeq2)
set.seed(787)
(dds <- makeExampleDESeqDataSet(n=5000, m=12, dispMeanRel = function(x) 4/x + 0.1))
#> class: DESeqDataSet
#> dim: 5000 12
```

```r
#> metadata(1): version
#> assays(1): counts
#> rownames(5000): gene1 gene2 ... gene4999 gene5000
#> rowData names(3): trueIntercept trueBeta trueDisp
#> colnames(12): sample1 sample2 ... sample11 sample12
#> colData names(1): condition
summary(rowMeans(counts(dds)))
#>    Min. 1st Qu.  Median    Mean 3rd Qu.    Max.
#>    0.00    6.25   16.00   40.94   39.85 1994.42
```

We need to normalize between the samples by estimating the DESeq2 size factors, and convert the *DESeqDataSet* object to *DGEList* object with equivalent normalization factors. This facilitates the comparison between DESeq2 and limma-voom pipeline.

```r
dge = DEFormats::as.DGEList(dds)
dds = estimateSizeFactors(dds)
dge$samples$norm.factors <- sizeFactors(dds) * mean(dge$samples$lib.size) / dge$samples$lib.size
dge$samples$norm.factors <- dge$samples$norm.factors / exp(mean(log(dge$samples$norm.factors)))
```

We first use the negative binomial modeling in DESeq2 to obtain gene-wise dispersion estimates and the mean-dispersion function.

```r
## dispersion estimates from DESeq2
#dds = estimateDispersionsGeneEst(dds)
#dds = estimateDispersionsFit(dds)
#dds = estimateDispersionsMAP(dds)
dds = estimateDispersions(dds) #equivalent to calling the above three steps
#> gene-wise dispersion estimates
#> mean-dispersion relationship
#> final dispersion estimates
## the DESeq2 estimates of mean-dispersion function
(mdfun <- dispersionFunction(dds))
#> function (q)
#> coefs[1] + coefs[2]/q
#> <bytecode: 0x1e1039e8>
#> <environment: 0x1c4bb458>
#> attr(,"coefficients")
#> asymptDisp  extraPois
#>  0.0938893  4.2555735
#> attr(,"fitType")
#> [1] "parametric"
#> attr(,"varLogDispEsts")
#> [1] 0.3138238
#> attr(,"dispPriorVar")
#> [1] 0.25
## the quantity that essentially determines the VST transformation (true value=(4+1)/0.1=50)
(attr(mdfun, 'coefficients')['extraPois'] + 1) / attr(mdfun, 'coefficients')['asymptDisp']
#> extraPois
#>  55.97628
```

As expected, the DESeq2 estimated mean-dispersion function parameters are close to the true values. We next show the proposed method to derive these estimates from limma-voom pipeline. We derive the gene-level dispersion estimates as follows:

```r
## dispersion estimates approximation from voom
design = model.matrix(~group, dge$samples)
vm = limma::voom(dge, design, plot = TRUE, save.plot=TRUE)
```

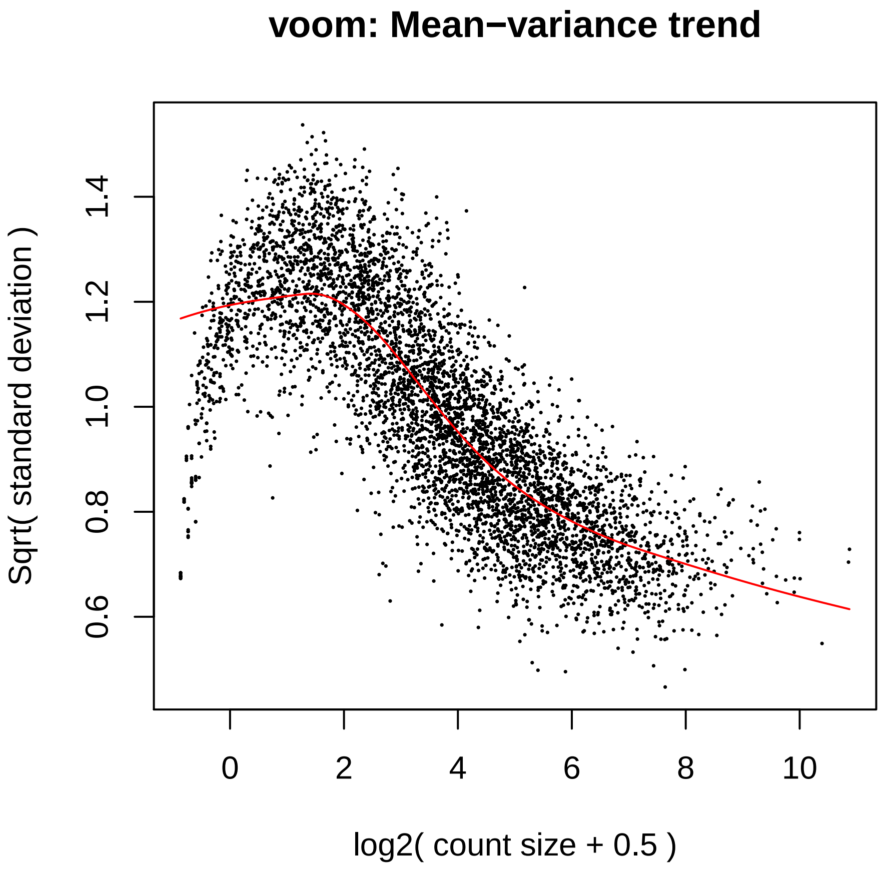


```r
md.xy = transfvoomxy(vm$voom.xy)
```

We then fit the mean-dispersion function and compare the estimates with truth:

```r
## filter out low quality dispersion estimates
table(good <- md.xy$y > 0 & md.xy$x > 10)
#>
#> FALSE  TRUE
#>  2104  2884
midx = match(names(md.xy$y), rownames(mcols(dds)))
## check gene-wise average count estimates
#summary(lm(VOOMmean ~ mean, data = data.frame(mean=mcols(dds)$baseMean[midx], VOOMmean=md.xy$x)))
## limma-voom based estimates of gene-wise dispersions versus truth
summary(lm(VOOMdispGeneEst ~ trueDisp, data = data.frame(trueDisp=mcols(dds)$trueDisp[midx],
        VOOMdispGeneEst=md.xy$y), subset=good))
#>
#> Call:
#> lm(formula = VOOMdispGeneEst ~ trueDisp, data = data.frame(trueDisp = mcols(dds)$trueDisp[midx],
#>     VOOMdispGeneEst = md.xy$y), subset = good)
#>
#> Residuals:
```

```
#>      Min       1Q   Median       3Q      Max 
#> -0.49650 -0.09685 -0.02919  0.06125  1.39233 
#> 
#> Coefficients:
#>             Estimate Std. Error t value Pr(>|t|)    
#> (Intercept)  0.01867    0.00909   2.054     0.04 *  
#> trueDisp     1.04811    0.03589  29.204   <2e-16 ***
#> ---
#> Signif. codes:  0 '***' 0.001 '**' 0.01 '*' 0.05 '.' 0.1 ' ' 1
#> 
#> Residual standard error: 0.1813 on 2882 degrees of freedom
#> Multiple R-squared:  0.2284, Adjusted R-squared:  0.2281 
#> F-statistic: 852.9 on 1 and 2882 DF,  p-value: < 2.2e-16
disp = cbind(as.data.frame(mcols(dds)[midx, c('trueDisp','dispGeneEst')]),
             VOOMdispGeneEst=md.xy$y)
cor(disp[good,])
#>                  trueDisp dispGeneEst VOOMdispGeneEst
#> trueDisp        1.0000000   0.5207215       0.4778618
#> dispGeneEst     0.5207215   1.0000000       0.9352717
#> VOOMdispGeneEst 0.4778618   0.9352717       1.0000000
## the limma-voom based estimates of mean-dispersion function
(mdfun2 <- DESeq2:::parametricDispersionFit(md.xy$x[good], md.xy$y[good]))
#> function (q) 
#> coefs[1] + coefs[2]/q
#> <bytecode: 0x1e1039e8>
#> <environment: 0x24256378>
#> attr(,"coefficients")
#> asymptDisp  extraPois 
#>  0.1056742  4.1441887
## the quantity that essentially determines the VST transformation (true value=(4+1)/0.1=50)
(attr(mdfun2, 'coefficients')['extraPois'] + 1) / attr(mdfun2, 'coefficients')['asymptDisp']
#> extraPois 
#>   48.6797
```

These results show the proposed method gives accurate estimates for the desired VST based on limma-voom pipeline.


### Appendix

```r
sessionInfo()
#> R version 3.5.0 (2018-04-23)
#> Platform: x86_64-pc-linux-gnu (64-bit)
#> Running under: CentOS release 6.10 (Final)
#>
#> Matrix products: default
#> BLAS: /app/R-3.5.0@i86-rhel6.0/lib64/R/lib/libRblas.so
#> LAPACK: /app/R-3.5.0@i86-rhel6.0/lib64/R/lib/libRlapack.so
#>
#> locale:
#>  [1] LC_CTYPE=en_US.UTF-8       LC_NUMERIC=C
#>  [3] LC_TIME=en_US.UTF-8        LC_COLLATE=en_US.UTF-8
#>  [5] LC_MONETARY=en_US.UTF-8    LC_MESSAGES=en_US.UTF-8
#>  [7] LC_PAPER=en_US.UTF-8       LC_NAME=C
#>  [9] LC_ADDRESS=C               LC_TELEPHONE=C
#> [11] LC_MEASUREMENT=en_US.UTF-8 LC_IDENTIFICATION=C
#>
#> attached base packages:
#> [1] parallel  stats4    stats     graphics  grDevices utils     datasets
#> [8] methods   base
#>
#> other attached packages:
#>  [1] DESeq2_1.21.16              SummarizedExperiment_1.12.0
#>  [3] DelayedArray_0.8.0          BiocParallel_1.15.15
#>  [5] matrixStats_0.54.0          Biobase_2.42.0
#>  [7] GenomicRanges_1.33.13       GenomeInfoDb_1.18.1
#>  [9] IRanges_2.16.0              S4Vectors_0.20.1
#> [11] BiocGenerics_0.28.0         voomst_0.1.0.0
#> [13] vimcom_1.2-8                setwidth_1.0-4
#> [15] colorout_0.9-9
#>
#> loaded via a namespace (and not attached):
#>  [1] edgeR_3.24.3           bit64_0.9-7            splines_3.5.0
#>  [4] Formula_1.2-3          assertthat_0.2.0       DEFormats_1.10.1
#>  [7] latticeExtra_0.6-28    blob_1.1.1             GenomeInfoDbData_1.2.0
#> [10] yaml_2.2.0             RSQLite_2.1.1          pillar_1.3.1
#> [13] backports_1.1.3        lattice_0.20-38        limma_3.38.3
#> [16] glue_1.3.1             digest_0.6.18          RColorBrewer_1.1-2
#> [19] XVector_0.22.0         checkmate_1.9.1        colorspace_1.4-0
#> [22] htmltools_0.3.6        Matrix_1.2-15          plyr_1.8.4
#> [25] XML_3.98-1.16          pkgconfig_2.0.2        genefilter_1.64.0
#> [28] zlibbioc_1.28.0        purrr_0.3.0            xtable_1.8-3
#> [31] scales_1.0.0           tibble_2.0.1           htmlTable_1.13.1
#> [34] annotate_1.60.0        ggplot2_3.1.0          nnet_7.3-12
#> [37] lazyeval_0.2.1         survival_2.43-3        magrittr_1.5
#> [40] crayon_1.3.4           memoise_1.1.0          evaluate_0.12
#> [43] foreign_0.8-71         tools_3.5.0            data.table_1.12.8
#> [46] stringr_1.3.1          locfit_1.5-9.1         munsell_0.5.0
#> [49] cluster_2.0.7-1        AnnotationDbi_1.44.0   bindrcpp_0.2.2
#> [52] compiler_3.5.0         rlang_0.3.1            grid_3.5.0
#> [55] RCurl_1.95-4.11        rstudioapi_0.9.0       htmlwidgets_1.3
#> [58] bitops_1.0-6           base64enc_0.1-3        rmarkdown_1.11
#> [61] gtable_0.2.0           DBI_1.0.0              R6_2.3.0
```

```
#> [64] gridExtra_2.3         knitr_1.21            dplyr_0.7.8
#> [67] bit_1.1-14            bindr_0.1.1           Hmisc_4.1-1
#> [70] stringi_1.5.3         Rcpp_1.0.0            geneplotter_1.60.0
#> [73] rpart_4.1-13          acepack_1.4.1         tidyselect_0.2.5
#> [76] xfun_0.4
```
